# The time- and space-varying roles of human mobility in shaping urban dengue epidemics

**DOI:** 10.1101/2025.06.03.25328731

**Authors:** Cathal Mills, Geziel dos Santos de Sousa, Antonio Silva Lima Neto, Vasco Furtado, Sen Pei, Moritz U.G. Kraemer, Christl A. Donnelly

## Abstract

Dengue outbreaks continue to cause large morbidity and mortality globally. Previous work has shown that human movement alongside weather, climatic, ecological, and socioeconomic factors influence outbreak size, persistence, and geographical spread. However, the relative importance of human movement is unclear for the establishment, rapid expansion, and persistence of dengue in urban settings. Relatedly, the extent to which neighbourhoods differentially influence outbreaks over time is unclear. To address these gaps, we developed a multi-model framework that integrates Bayesian hierarchical modelling and data-driven, deep-learning-based approaches to parameter inference and out-of-sample probabilistic predictions of dengue in Fortaleza, Brazil. We apply our framework to dengue outbreaks at the neighbourhood level, using epidemiological surveillance data alongside public transportation data. We use information criteria, scoring rules, and explainability metrics to measure the models’ performance and ultimately explain how human mobility shapes disease dynamics. We find that human mobility is a consistent driver of dengue outbreaks over the five-year period studied. We also find that human movement is a more important driver of transmission dynamics between neighbourhoods than transmission within them, both within and across dengue seasons, and that spatially, the relative importance of different neighbourhoods to transmission elsewhere is relatively constant over time. Our framework highlights the time- and space-varying roles of human mobility and is applicable to outbreaks of other infectious diseases and to other questions of relative epidemic drivers.

## 1 Introduction

Dengue is a vector-borne disease caused by the dengue virus (DENV) that has four distinct serotypes and is spread by *Aedes* mosquitoes, transmitted between humans primarily by *Ae. aegypti* and to a lesser extent, *Ae. albopictus*. Dengue is the most rapidly expanding mosquito-borne disease, approximately half of the world’s population is currently at risk of infection^[1,2]^, and the incidence and distribution are expanding^[3]^. Faced with the growing public health challenges associated with severe dengue, there has been an increasing effort to improve our understanding of the biological, human, and environmental influences on dengue epidemics around the world. Modelling is one way to provide such understanding, and a wide range of approaches have been used, including statistical, mechanistic (or mathematical), machine learning (ML), and deep learning frameworks. Given the climatic sensitivities across both the pathogen (DENV) and mosquito vector[4,5,6], a common research objective has been to quantify how transmission dynamics have been shaped by recent climatic conditions and their interactions with human (e.g. urbanisation, human mobility, and socioeconomic factors[7,8,9]) and epidemiological influences (e.g. serotype replacement, serotype competition, and population immunity[10,11]).

In recent years, human movement has been identified as an important driver in dengue epidemic dynamics. In Pakistan, a mechanistic modelling study used mobile phone data and climate data to reveal that the inclusion of human mobility improved predictions of the spread and timing of dengue epidemics^[12]^. Mechanistic approaches have similarly been used in southern China and Singapore to model dengue spread within and across cities ^[13,14,15,16]^. Statistical frameworks, such as Bayesian hierarchical models, have generally incorporated the effects of human mobility using either human movement matrices, distance-based approximations, or neighbourhood-based approximations^[8,17]^. In the city of Fortaleza in Brazil, both mobility-informed artificial neural networks and mechanistic models were employed at a fine spatial resolution (neighbourhood level). The inclusion of human mobility (bus transportation) data again yielded improvements in prediction accuracy^[18]^. A separate predictive modelling study^[19]^ used data from the COVID-19 pandemic to identify human mobility as a crucial factor in shaping dengue epidemic dynamics in Fortaleza. At a longer-term horizon, results from Harish et al. ^[20]^, based on a hierarchical model of geographical expansion, indicated important synergistic roles for short- and long-distance human movement, alongside suitable environmental conditions, for the geographical expansion of dengue in Brazil and Mexico.

However, while the increasing availability of mobility data (in different guises) has enabled such modelling studies approaches (which extend beyond mobility approximations^[21]^), we still have a poor understanding of how the relative importance of human movement in dengue outbreaks varies, within and across seasons and across space. Such time-varying effects may be important, as genomic studies indicate that once dengue is established, reimportations remain important^[22]^. We consider here when, where, and how human movement shapes urban dengue outbreaks at the neighbourhood level in a city (Fortaleza, Brazil) where dengue is endemic for over 30 years. Research questions we ask are: In an urban context, how does the role of human mobility change through time? How does human mobility shape the epidemic dynamics once the outbreak is under way? How does the relative importance of human mobility change across years? How do neighbourhoods differentially impact the city-wide dynamics of dengue over time? We investigate these questions using mobility-informed statistical and deep learning models, alongside statistically principled multi-model development and new evaluation procedures. Our methodology for mobility-informed modelling allows us to infer the high-resolution time- and space-varying relative importance of human mobility for driving urban dengue outbreak dynamics, and quantify the impacts of human mobility on source-sink dynamics of neighbourhoods. Looking ahead, our framework can provide the insights necessary for public health authorities to design more targeted and optimally timed public health interventions, both for dengue and other infectious diseases.

## 2 Materials and methods

### 2.1 Study area and data

Fortaleza is a coastal city in north-east Brazil with a population of approximately 2.4 million people^[23]^. It is the fifth-largest city in Brazil and consists of 119 neighbourhoods/bairros (Figure 1A).

**Figure 1:**
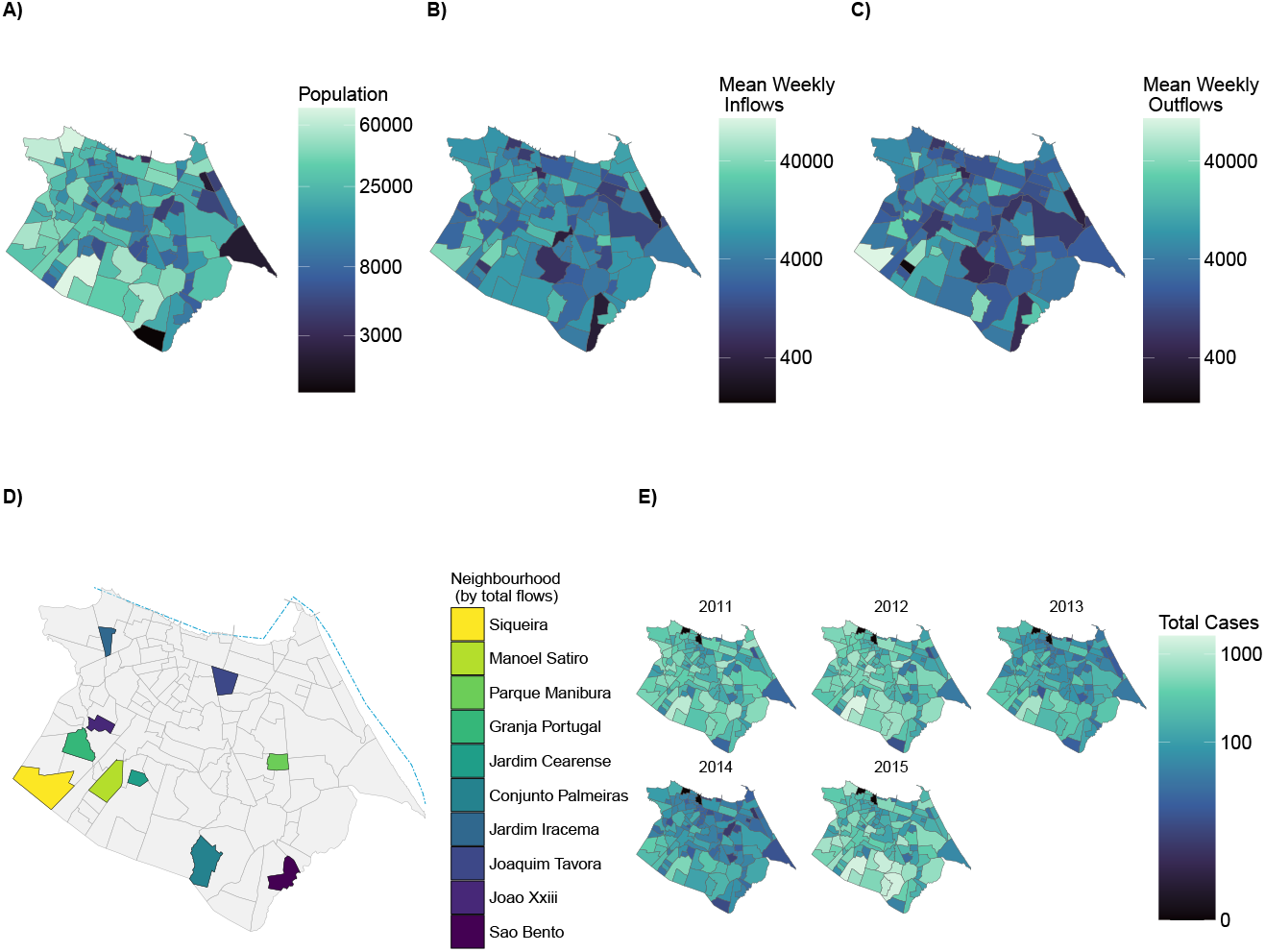
Demography, human movement, and dengue cases in Fortaleza, Brazil. Fortaleza is a coastal city in north-east Brazil, with a population of approximately 2.4 million people[23]. Within Fortaleza, there are 119 neighbourhoods (known as bairros), and all our data and analyses are at the neighbourhood level. **A)** The population (visualised on a logarithmic scale) of each of the 119 neighbourhoods, **B)** and **C)**: The mean weekly total number (visualised on a logarithmic scale) of inflows and outflows respectively for each of the 119 neighbourhoods. **D)** The neighbourhoods with the top 10 total number of inflows and outflows (visualised on a logarithmic scale) across all 52 weeks. Neighbourhoods are ordered in descending order from highest to lowest flows, and the blue line visualises the coastline of Fortaleza. **E)** The cumulative reported dengue cases (visualised on a logarithmic scale) in each neighbourhood per year of the testing period (2011–2015). The time series of cases within each year is visualised Figure 3A–B, while a summary of cases per week are visualised in Figure SI 3.

Dengue is endemic in Fortaleza. The first recorded dengue outbreak occurred in 1986 and was caused by the introduction of DENV-1. DENV-2 was first introduced into Fortaleza in 1992, with the first major epidemic occurring in 1994 when approximately 660,000 dengue cases were reported in Fortaleza (approximately 84% of all cases in Brazil) driven in part by the suspension of mosquito control activities in late 1993 due to a cholera outbreak^[24,25]^. By 2012, all four serotypes of DENV had been detected in the city. Female *Ae. aegypti* are the primary vector of DENV transmission across Brazil. Suitable conditions, for the development of their larval habitat and DENV transmission, are provided by the highly urbanised Fortaleza neighbourhoods and the equatorial savannah climate (with distinct tropical rainy and dry seasons)^[7,15,26]^.

To measure human mobility, we used previously published data for the public bus transportation system in Fortaleza; the most widely used mode of public transport with an estimated 700,00 people using the system every day^[27,28,29]^. The mobility data were available on a weekly basis for 2015. Using the coordinates of the origin and destination location, we obtained a directed measure of the number of flows 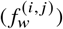 from neighbourhood *i* to neighbourhood *j* for week *w*. In the absence of further available data, we assumed the weekly mobility matrix (i.e the directed adjacency matrix) from 2015 applied to corresponding weeks in all other years (2007–2014). The mobility data were described extensively in previous work^[18,27,28,29]^.

We sourced data on newly reported dengue cases from[18], which used reported cases from the Sistema de Monitoramento Diário de Agravos—SIMDA (Daily Disease Occurrence Monitoring System). These cases arise from diagnoses in hospitals or primary health care units. Note that dengue cases in Brazil can be confirmed solely on the basis of clinical and epidemiological criteria, without laboratory tests. The data were reported on a weekly basis (using diagnosis dates) for each of the 119 neighbourhoods according to the cases’ home addresses.

For meteorological variables, we calculated daily mean temperature (at a 2 metre level) and daily total precipitation for Fortaleza using data from the ERA5-Land reanalysis dataset[30]. Due to data resolution, the same climatic conditions were used for each neighbourhood, and are generally characterised by year-round stable, suitable temperatures for dengue transmission and distinct rainy and dry seasons (Figure SI 1).

We sourced data on newly reported dengue cases from^[18]^, which used reported cases from the Sistema de Monitoramento Diário de Agravos—SIMDA (Daily Disease Occurrence Monitoring System). These cases arise from diagnoses in hospitals or primary health care units. Note that dengue cases in Brazil can be confirmed solely on the basis of clinical and epidemiological criteria, without laboratory tests. The data were reported on a weekly basis (using diagnosis dates) for each of the 119 neighbourhoods according to the cases’ home addresses.

For meteorological variables, we calculated daily mean temperature (at a 2 metre level) and daily total precipitation for Fortaleza using data from the ERA5-Land reanalysis dataset^[30]^. Due to data resolution, the same climatic conditions were used for each neighbourhood, and are generally characterised by year-round stable, suitable temperatures for dengue transmission and distinct rainy and dry seasons (Figure SI 1).

### 2.2 Model development and fitting periods

To guard against optimistic bias and overfitting, we used data from 2007 to 2010 inclusive as a model development period. During this period, we selected combinations of covariates, developed model structures, and made other model choices based on information criteria (see below).

We then focused on data from 2011 to 2015 inclusive to test model performance and draw insights. Rolling windows allowed us to estimate spatiotemporal variations in mobility effects without explicitly modelling the time variation for each observation, helping to guard against non-identifiability issues which could be caused by time-varying effects for each time point and neighbourhood.

After exploratory analyses (2007 to 2010), we chose a 17-week (approximately four-month) rolling window to allow sufficient learning of parametric relationships in our Bayesian hierarchical model, without losing excessive temporal resolution. The model learned about mobility influences within each rolling window, a procedure which also prevents averaging over potential data distribution shifts (or equivalently, changing mobility-incidence-rate relationships). For our deep learning models, we fitted these models using all information up to a given time *t*, due to the need for larger volumes of data to estimate covariate effects (e.g., mobility influences). Unless otherwise stated, all evaluation occurs in the rolling windows.

### 2.3 Bayesian hierarchical models

We developed Bayesian hierarchical approaches to modelling *y*_*it*_, the reported neighbourhood-level (*i*) weekly (*t*) case counts of dengue. We used a Negative Binomial (NegBin) likelihood to allow for potential overdispersion in the case data.

We incorporated the graph structures (from the weekly mobility data) using a lagged mobility-weighted sum of all other neighbourhoods’ past dengue incidence rates (DIRs) per 100,000, which provides an estimated force of dengue importations. This approach was a computationally cheap (albeit limited, see Discussion) way to incorporate spatiotemporal structure that was informed by human mobility data. We used mobility flows and (reported) DIR from the previous week as inflow of infected individuals is less likely to result in new reported human cases within the reporting week (given the intermediate role of the mosquito vector, viral replication rates within the host and vector, and possible reporting delays).

We developed and fitted a range of candidate models (Supplementary Material A.2) in R version 4.4.0 using the cmdstanr interface to Stan, a platform for Bayesian inference^[31,32,33]^. Using the R package loo, we estimated the out-of-sample generalisability of each model using ELPD-LOO; leave-one-out (LOO) estimates of the expected log pointwise predictive density (ELPD)^[34]^. Based on results for the model development period, we selected the following candidate model which assumes a fixed mobility parameter (across space) and a space-varying autoregressive parameter:

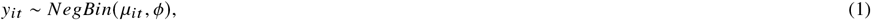

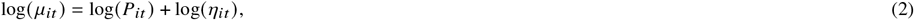

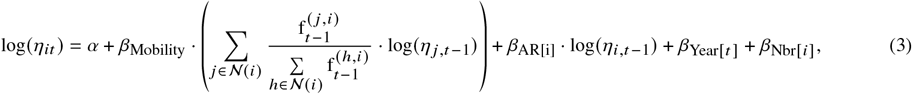

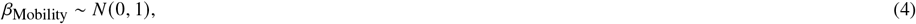

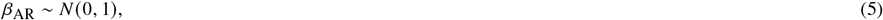

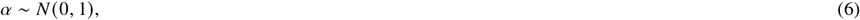

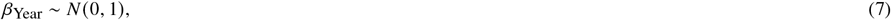

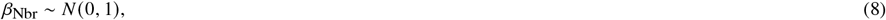

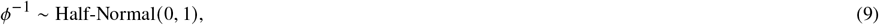

where *μ*_*it*_ denotes the mean parameter of the Negative Binomial distribution and *ϕ* is the reciprocal overdispersion parameter. We used a population offset log *P*_*it*_ such that *η*_*it*_ represented the DIR per 100,000 population. We then modelled the (logarithm of) DIR with i) an intercept *α*, ii) an estimated force of dengue importations – a mobility effect controlled by fixed parameter *β*_Mobility_ and the lagged (logarithm of) DIR of all other neighbourhoods, 𝒩(*i*), weighted by the proportion of lagged inflows from such other neighbourhoods 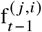, iii) a potentially non-stationary and neighbourhood-specific autoregressive process of order one, controlled by *β*_AR[i]_ (which allows for within-neighbourhood transmission caused by factors beyond mobility), iv) a year-to-year random effect (i.e. *β*_Year_ is a varying intercept for each year), and v) a neighbourhood-specific random effect (i.e. *β*_Nbr_ is a varying intercept for each neighbourhood). Modelling the effects on a DIR scale allowed us to disentangle each of these effects from population artefacts (e.g. large versus small neighbourhoods). Our goal was to understand the relative importance of human-mobility-driven transmission versus local transmission and the mobility-driven roles of different neighbourhoods in epidemic spread. We also explored explicitly accounting for climatic drivers (weekly total precipitation and average temperature), but the lack of neighbourhood-specific data meant that, causally, climatic covariates could not provide information for climate-driven local transmission dynamics (see Discussion).

A fixed parameter (within each window) for mobility does not assume that the effects of mobility (on each neighbourhood’s DIR) are homogeneous across space and time, as i) both the human movement and human case data vary across space and time, and ii) the model can be fitted in rolling windows (which allows spatiotemporal variations in the autoregressive term and the levels at which inflows of infected individuals generate new infections).

#### 2.3.1 Mobility effects across space and over time

Using this model (eqs (1)–(9)), we inferred parameter-based estimates of i) how human mobility varies in terms of its relative importance within and across neighbourhoods over time, and ii) how human mobility impacts the time-varying, relative contributions (i.e. source-sink dynamics) that neighbourhoods make to the epidemics of other neighbourhoods.

We computed the following ratio, *r*_*i*_ (*t*), for each neighbourhood *i* at time *t*:

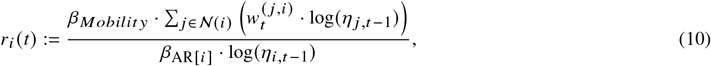

Where 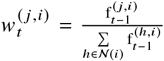 is the weight assigned to neighbourhood *i* for the estimated inflows of infected travellers from neighbourhood *j*.

Eq. (10) measures the estimated time-varying relative importance of human mobility; contributions (to neighbourhood *i*’s epidemic) from all other neighbourhoods, relative to its own autoregressive epidemic process, with values of *r*_*i*_ *(t*) greater (less) than 1 indicating a greater (lower) relative importance of mobility-driven case importations. Using DIRs enabled comparison of the relative importance of human mobility across neighbourhoods of potentially different sizes. We compared how *r*_*i*_ *(t)* differed across space (*i*) and over time (*t*), which helped us investigate the question of whether mobility’s relative importance in driving dengue seasons is temporally homogeneous across space – i.e. does the importance follow similar patterns within and across seasons in different neighbourhoods?

We also estimated source-sink dynamics by deriving formulae for 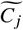. This variable was standardised per window (across neighbourhoods) and reflects the relative contributions of a given neighbourhood *j* to the dengue epidemic dynamics in all other neighbourhoods. Values of 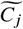 greater (less) than 0 indicate that a given neighbourhood has contributed more (less) to transmission elsewhere, relative to the mean contribution of neighbourhoods.

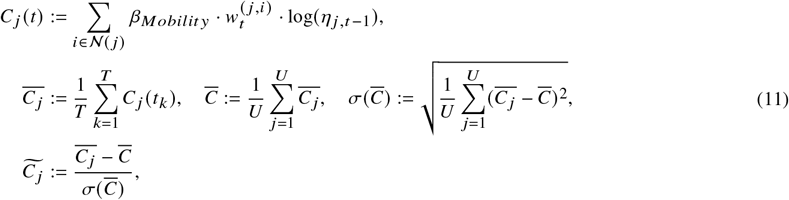

where *T* is the length of the rolling window (i.e. the 17-week period to which the model was fitted), *U* is the number (119) of neighbourhoods, |𝒩 (*j*) | = *U* − 1 is the number of neighbours in 𝒩 (*j*), and 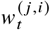 is the weight assigned for the proportion of outflows that leave neighbourhood *j* to visit neighbourhood *i*. Then, by tracking 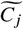 in each rolling window, we measured how the relative contribution of a single neighbourhood *j* (to all other neighbourhoods) varies over time. We standardised per window to quantify *relative* contributions (source/sink) of neighbourhoods during different stages of dengue seasons (i.e. accounting for different epidemic magnitudes in different windows).

We assessed the importance and roles of human mobility via these derived estimates (Eqs (10) – (11)), alongside information criteria (ELPD LOO) and out-of-sample predictive performance for one-step-ahead forecasts using the Weighted Interval Score (WIS)^[35]^ (a proper scoring rule which balances sharpness and calibration, see below).

### 2.4 Data-driven, deep learning models

We also developed data-driven, deep learning models (denoted as *DL models* in visualisations). The objective was to draw more robust conclusions by leaning on different approaches to modelling the effects of human mobility.

We implemented two foundational time series models (with and without human mobility effects), both based on TimeGPT; a transformer-based, foundational time series model that was pre-trained on more than 100 billion data points from a range of domains^[36,37]^. In both models, we modelled the logarithm of incident cases *y*_*it*_ in neighbourhood *i* at time *t* as our response for two reasons; i) we constrained our model’s predictions to be positive, and ii) we measured one-step-ahead probabilistic forecasts using a recommended logarithmic scale^[38]^. In only the mobility-informed deep learning model, we used a covariate of a mobility-weighted sum of other neighbourhoods’ past cases *m*_*i,t*_ (on a logarithmic scale), which entered via

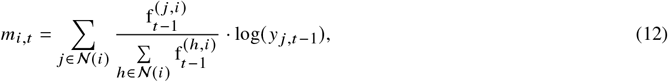

where 𝒩(*i*) and 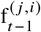 are defined as before. In both models (with and without the inclusion of human mobility effects), we further included the neighbourhood as a factor variable (i.e. a spatial effect) and the neighbourhood’s most recently observed cases log *y*_*i,t* 1_ (on a logarithmic scale, i.e. similar to an autoregressive covariate). Unless otherwise stated, we use the mobility-informed model when discussing the deep learning model’s results.

These foundational time series models facilitate probabilistic predictions (via quantile regression with conformal inference) in the form of predictive quantiles of the forecast/predictive distribution. Therefore, to ensure comparability when assessing models’ probabilistic predictions, we used the WIS (via the scoringutils R package^[39]^), which is an approximation to the Continuous Ranked Probability Score (CRPS) for quantile-based forecasts, which we represented using 23 predictive quantiles, as in previous forecasting work^[40,41]^.

A key challenge of any deep learning model is explainability^[42]^. To understand why our deep learning models made its *individual predictions*, we used SHAP (SHapley Additive exPlanations) values. These improve upon feature importance metrics which can only measure overall importance to the model and cannot measure the directional influences of each feature. In contrast, SHAP values, based on a concept from game theory, measure the contribution of each feature/covariate to the model’s individual predictions^[43]^.

We denote 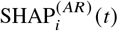 and 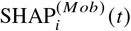 as the SHAP values (i.e. estimated contributions) of the autoregressive influence, log (*y*_*i,t* 1_) and mobility-weighted inflows, *m*_*it*_ to the model’s forecast (of the logarithm of cases) for neighbourhood *i* and time *t*. We can then derive an estimate for the relative importance of human mobility that is analogous to *r*_*i*_ (*t*) (Eq (10)) from the Bayesian hierarchical model:

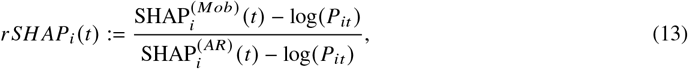

where *P*_*it*_ is defined as before as the population offset (population divided by 100,000) needed to convert to (the logarithm of) DIR scale.

This formulation, eq. (13), again, can measure the *estimated* relative importance of human mobility in individual neighbourhoods with potentially different population sizes. Of course, we caveat our approach with the causal limitation that SHAP values measure feature contributions to the models’ individual forecast. There is no explainability metric that can guarantee a correct causal result. Although our multi-model, triangulation-like approach^[44]^ may mitigate this causal limitation, recent research has demonstrated causal misalignments when using SHAP values for deep learning models in epidemiological settings (even when the models are well-calibrated and highly predictive)^[45]^. In our setting, a SHAP value does not measure the contribution to observed case data nor to the true latent infection process. Furthermore, unlike our Bayesian hierarchical models which are generative and more mechanistic, the SHAP and rSHAP values of our deep learning models are not constrained by even a semi-mechanistic structure. For example, the SHAP value for the mobility-weighted influence 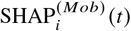 may be positive when *m*_*it*_ = 0 and could even be negative (when indicating that the base forecast should be decreased when *m*_*it*_ takes a particular, e.g. low, value). It is also possible that another unmodelled factor beyond mobility drives the difference between the SHAP values.

### 2.5 Evidence synthesis

Using both modelling approaches (Bayesian and deep learning), we generated one-step-ahead probabilistic forecasts of dengue cases (on a logarithmic scale, as recommended for epidemic forecasts^[38]^), and measured out-of-sample predictive performance of the mobility-informed models versus i) their analogous models with the mobility effect omitted and ii) a naive, baseline forecasting model^[41]^. This one-step-ahead probability distribution is equivalent to the predictive likelihood. Improvements over reference models, as measured by relative WIS, rWIS (WIS for given model divided by WIS for an analogous model without mobility or baseline model), for different rolling time windows and neighbourhoods, allowed us to quantify the extent to which mobility-driven importations were important for generating the observed incidence data. For the Bayesian hierarchical model, we also measured improvements (upon the analogous model without mobility) using LOO estimates of the ELPD, thus measuring how much mobility effects improve (estimated) out-of-sample generalisability.

We interpret *r*_*i*_ *(t*) and *r SH AP*_*i*_ *(t*) from our Bayesian hierarchical model and deep learning model as proxies for the relative importance of human mobility over time in driving epidemics. We compared values for *r*_*i*_ *(t*) and *r SH AP*_*i*_ *(t*) using Pearson’s^[46]^ and Spearman’s correlation^[47]^, categorisation into binary and four-level categories, and t-tests with p-values (Bonferroni-adjusted^[48]^ when comparing multiple time series across neighbourhoods). Drawing comparison between these quantities ensured that we did not over-inflate the insights from a single modelling approach about the relative importance of human movement.

To understand the likely mechanisms behind values for *r*_*i*_ *(t*) and *r SH AP*_*i*_ *t*, we visualised relationships with factors such as epidemic stage (measured by the cumulative fraction of annual cases observed by a given week), year, and neighbourhood. We further developed simple generalised additive models (GAMs) for *r*_*i*_ *t* and *r SH AP*_*i*_ *t*, consisting of a Gaussian likelihood with covariates of neighbourhood-specific smooth functions (thin-plate splines) for epidemic stage, annual temporal and neighbourhood-specific spatial random effects (to account for e.g. varying reporting rates), and the most recent observed cases (on a logarithmic) scale, *y*_*i,t* 1_. We compared alternative GAM models using the minimised generalised cross-validation score (GCV)^[49]^.

Due to its generative structure, the Bayesian hierarchical model permits estimates of how an individual neighbourhood *j* (via journeys of its infected residents/commuters) impact the epidemics in other neighbourhoods, as measured by 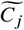.We quantified possible causes behind the values of 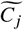 in different rolling windows using a similar procedure; visualisations and local contextualisation complemented by follow-up GAMs chosen based on their GCV. The candidate GAMs here are, by construction of 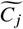,conceptually simpler as 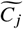 is a standardised per-rolling-window quantity, and so temporal effects (for e.g. annual reporting rates) are likely less relevant.

After model fitting, using Pearson’s correlation coefficients^[46]^, we also performed a simple correlation analysis of the full DIR time series (2007–2015) for the 119 neighbourhoods to understand the spatial synchrony across dengue seasons. Finally, to better understand the structure of the mobility network, for each week, we used i) modularity from the Louvain algorithm^[50]^ (a community detection algorithm that works by hierarchical clustering), ii) modularity from the Infomap algorithm^[51]^ (a community detection algorithm for directed, weighted graphs), and iii) Shannon’s entropy^[52]^ of the mobility flow matrix. Higher values of modularity and lower values of entropy indicate strong clustering (i.e. low mixing) and high concentration of flows in particular locations.

## 3 Results

The 119 neighbourhoods of Fortaleza have different population sizes (ranging from 1,342 to 76,044, Figure 1A) and therefore exhibit different human movement patterns (as approximated by the bus transportation data, Figure 1B–D) and consequently, may have different daytime (or ambient) population sizes due to inflows or outflows caused by commuter journeys in the city. In general, pairwise total flows between neighbourhoods were similar throughout the year (Figure SI 2), and there was a strong relationship (Pearson’s correlation coefficient *r* = 0.84, p-value *<* 0.0001) between a neighbourhoods’ total inflows and total outflows.

Across the studied period (2007–2015), the most severe annual dengue seasons were observed in 2008, 2011, 2012, and 2015 (Figures 1E and SI 3B), with peaks occurring almost across all neighbourhoods at similar times (Figures SI 3B, 3A). We visualised (Figure SI 3A) and quantified this high spatial similarity of time series, reflective of epidemic synchrony, using pairwise Pearson’s correlation coefficients^[46]^ between individual neighbourhoods (Median *r* = 0.74, Q1 = 0.66, Q3 = 0.81).

On their own, these exploratory results suggest, but do not confirm, roles for human mobility in driving dengue epidemics in Fortaleza. It is plausible that the highly synchronous dengue seasons across pairs of neighbourhoods (Figure SI 3A) are caused by single initial seeding events (as infectious hosts move from one neighbourhood to another) and subsequent similar epidemic trajectories.

### 3.1 Mobility consistently drives urban dengue outbreaks

Across our statistical and deep learning models, the inclusion of the effects of an estimated force of dengue importations (via weighted estimates of inflows, see Materials and methods) almost consistently improved model performance, as measured by probabilistic forecast performance metrics (rWIS, Figure 2A-C) and information criteria (ELPD LOO, Figure 2D). We observed these trends when we compared mobility-informed models to our baseline model which propagates the most recent observed cases as the next (median) forecast, and to analogous models which omitted the effects of the estimated force of dengue importations (Figures 2D and 3G). These trends were generally consistent i) across the 119 neighbourhoods, including in boundary neighbourhoods along the south and west of the city where our restriction to city-level mobility data could induce boundary effects (Figure 2B) and ii) over time, including during both peaks and troughs of dengue cases (Figure 2A and C). The only locations where we observed any predictive improvement from using the baseline model versus the mobility-informed Bayesian model were in Jacarecanga and Cristo Redentor; coastal/beach neighbourhoods which reported zero total dengue cases (Figure 1E). We found largely similar outperformance for the mobility-informed deep learning model (in 116 of 119 locations, Figure 2B). As a further model validation, the more generative (or semi-mechanistic) mobility-informed Bayesian hierarchical model consistently outperformed (within and across all dengue seasons) the corresponding data-driven deep learning model in terms of probabilistic predictive performance (WIS), suggesting greater out-of-sample model generalisability (Figure 1A and C).

**Figure 2:**
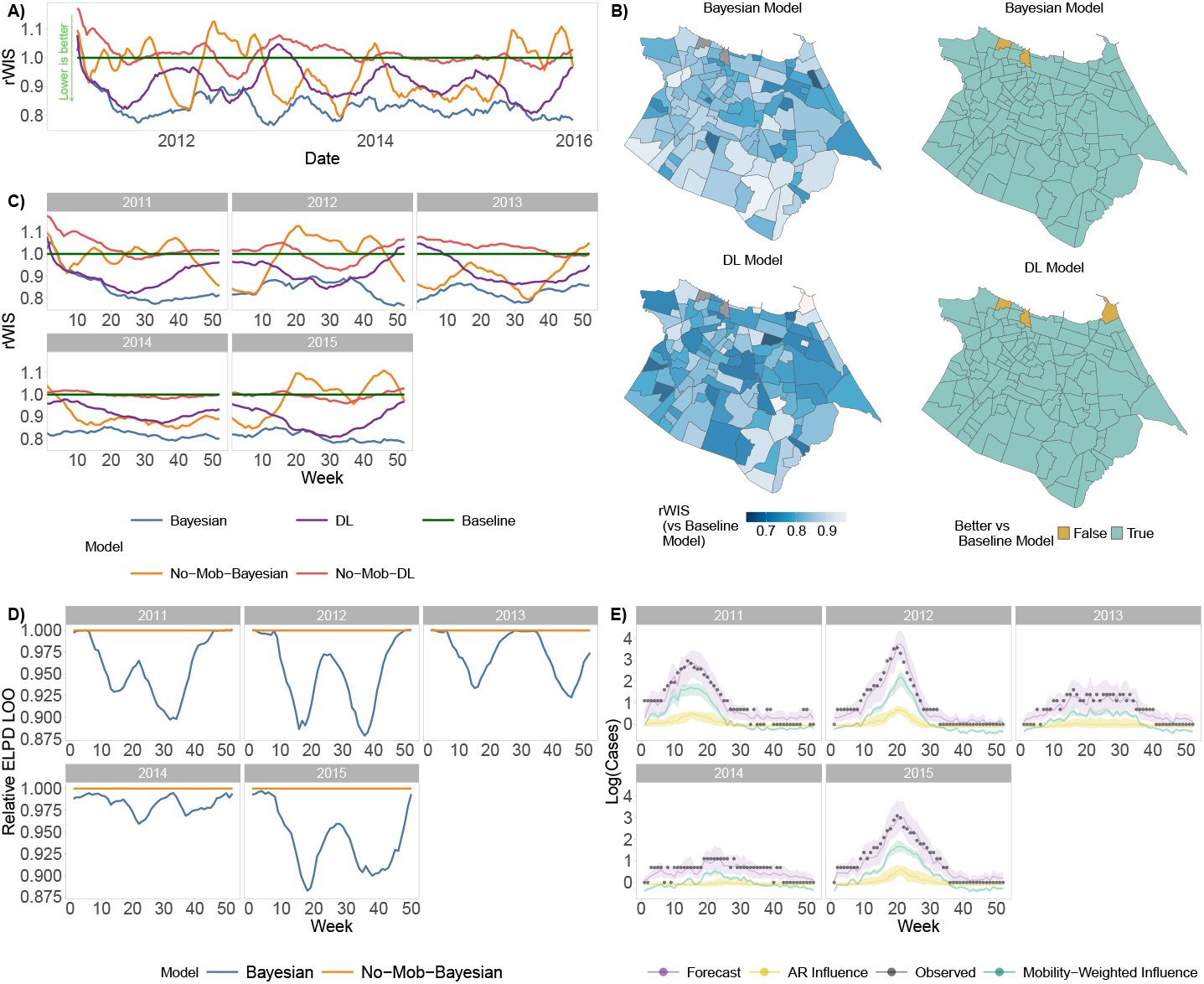
Capturing human mobility effects improves model performance. In approximate four-month trailing windows (17 weeks) of the testing period (2011-2015 inclusive), we visualise model performance for each of our four models across Bayesian hierarchical and deep learning (DL) approaches and compare performance to an epidemiologically naive baseline model (see Materials and methods). Any date shown represents the maximum date within the trailing window. **A)** Relative weighted interval score (rWIS) over time for each of the four models, where rWIS was calculated (relative to a baseline model, see Section 2.5) out of sample across all neighbourhoods. Lower values of the rWIS indicate superior predictive performance for out-of-sample case data (on a logarithmic scale). **B)** rWIS values for each of the 119 neighbourhoods across all trailing windows and a binary classification of whether the Bayesian/DL model outperformed the baseline (i.e. rWIS < 1). The grey shaded neighbourhoods on the north coastline are Jacarecanga and Cristo Redentor; neighbourhoods which reported zero total dengue cases across all seasons. **C)** Analogous to A), rWIS over time but using separate facets to capture model performance within each dengue season. Information criteria results for mobility-informed Bayesian hierarchical model. Higher ELPD LOO values indicate superior model generalisability. However, for visualisation of relative model performance over time (irrespective of the magnitude of observed data), we derived relative ELPD LOO values by dividing the ELPD LOO values for the Bayesian model by those for the analogous model without mobility (No-Mob-Bayesian). This was possible, as all ELPD LOO values were negative (across all windows and both models). Therefore lower values of this relative ELPD LOO indicate superior estimated model generalisability. Interquartile ranges of observed and forecasted cases (on a logarithmic scale) alongside the SHAP contributions from the neighbourhood-level autoregressive (AR) process and the mobility-weighted influence of the neighbours. Higher values of the SHAP values indicate greater contributions to the forecasted value.

### 3.2 Relative importance of human mobility over time to individual neighbourhoods

How the relative importance of human mobility changes throughout dengue outbreaks in urban settings remains poorly understood. We hinted above that human movement could play its most important role at the early stages of a dengue season (when seeding events can occur as a result of between-neighbourhood movement). Thereafter, human mobility could play less important and potentially negligible roles in sustaining transmission, once dengue is circulating widely in neighbourhoods of the city.

Using our Bayesian model, eqs (1)–(9), we assessed relative importance of human mobility for each neighbourhood via our estimates of *r*_*i*_ *(t*) – the ratio of (mobility-informed) case importations versus the neighbourhood’s own autoregressive generation of new cases from past cases. Temporally, we estimated different trends in the relative importance of human mobility in different dengue seasons across the individual neighbourhoods (Figure 3A-C, Figure SI 5). While the rWIS values indicated spatially consistent (Figure 3G) and temporally consistent (Figure 2A,C, and D) outperformance of the mobility-informed model relative to the analogous model without human mobility, the estimates of *r*_*i*_ *(t*) were spatially inconsistent within the annual dengue seasons (Figure SI 5C and Figure SI 6). As *r*_*i*_ *(t*) varied substantially across space and time, we used both a binary classification (Figure 3C) and a four-level classification (Figure 3E) of *r*_*i*_ *(t*) to further investigate the reasons/mechanisms behind variations in *r*_*i*_ *(t*). Across all observations (all neighbourhoods and time points), the times of estimated greater relative importance of human mobility (*r*_*i*_ *t*) most frequently coincided with times near the midpoint (Figure 3D) or epidemic peak (Figure SI 5, SI 6) of an annual dengue season. Despite these general observations, there is no distinct neighbourhood-level evidence of such systematic pattern.

**Figure 3:**
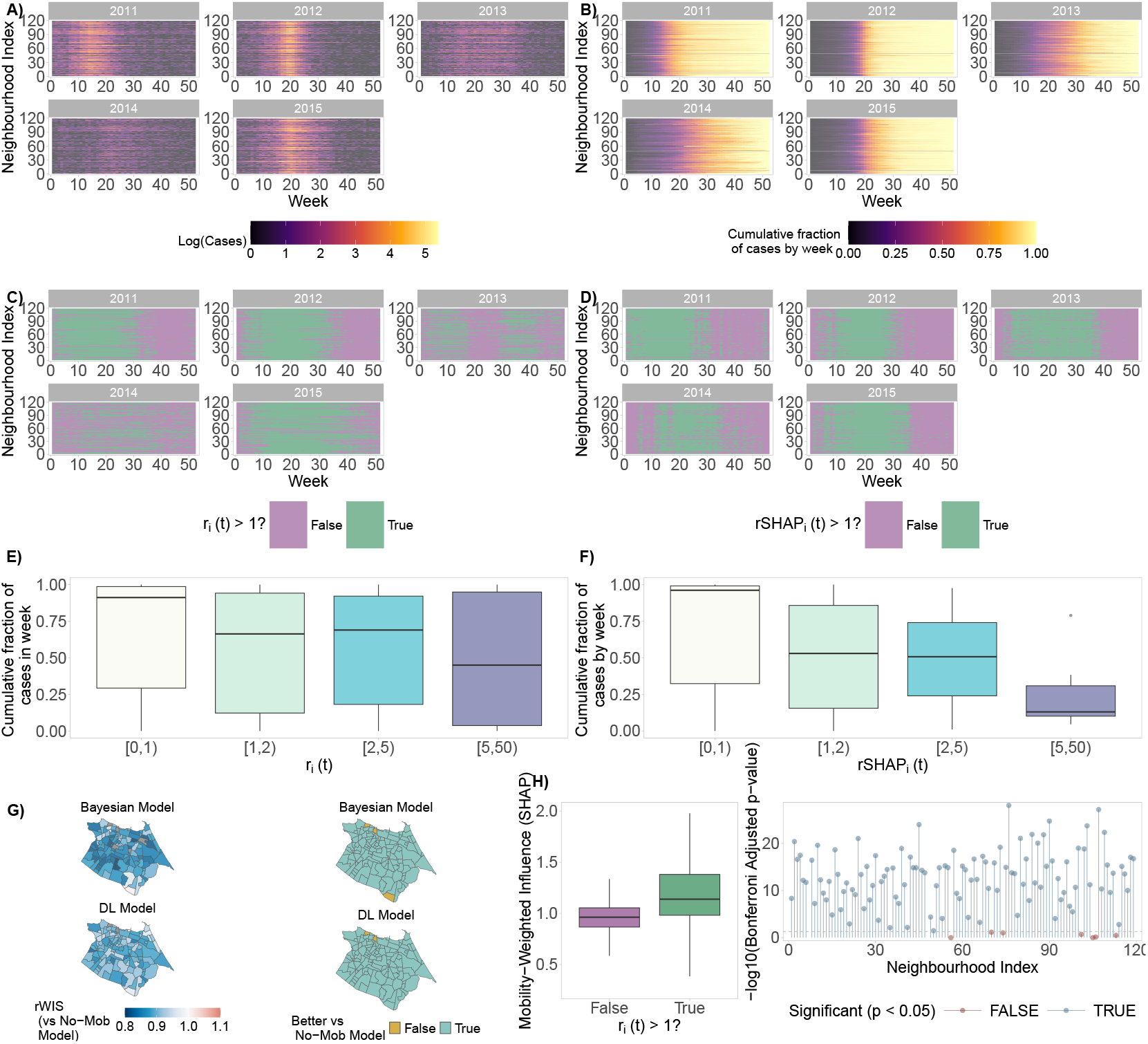
The time-varying relative importance of human mobility. Across 2011–2015 and the 119 neighbourhoods, we visualise the estimated relative importance of human-movement-based importations of dengue cases, versus the neighbourhoods’ own internal generation of new dengue cases. **A)** and **B)** visualise the observed cases (on a logarithmic scale) and cumulative fraction of annual cases observed for individual weeks across 2011-2015. Neighbourhoods are ordered in ascending order by their total annual number of flows (inflows and outflows). **C)** and **D** show the estimated relative importance from our Bayesian hierarchical and deep learning models respectively, where values of *r*_*i*_) and *r*SHAP_*i*_ *(t*) greater than one indicate greater relative importance of the case importations (see Materials and methods). Figure SI 5C-D is the analogous continuous version of these estimates of *r*_*i*_ *(t*) and *r*SHAP_*i*_ *(t*). **E)** and **F)** visualise the relationship between dengue season stage (measured by the cumulative fraction of cases) versus the values of *r*_*i*_ *t* and *r*SHAP_*i*_ *t*. **E)** Across the 119 neighbourhoods, the relative predictive performance of the Bayesian and deep learning model is visualised, using the relative weighted interval score (rWIS) compared against the corresponding models without a mobility effect. **G)** and **H)** capture the relationship between the model-based relative importance estimates *r*SHAP_*i*_ *(t*) and a binary version of *r*_*i*_ *(t*) (= 1 if *>* 1) using a boxplot and visualisation of the Bonferroni-adjusted p-values^[48]^ from a one-sided t-test of whether *r*SHAP_*i*_ *(t*)were significantly greater when *r*_*i*_ *(t*) *>* 1. The box and whiskers of each boxplot cover the interquartile range.

To test the robustness of model-based conclusions, we assessed the relationship between the Bayesian and deep learning models’ findings. First, we observed the same spatial and temporal outperformance of the predictions for the mobility-informed deep learning models, relative to analogous models without the mobility effect (as per rWIS, see Figure 2A-C and Figure 3G). We also compared the Bayesian-model-derived estimates *r*_*i*_ *(t*) with those from the SHAP values for the mobility-weighted influence, 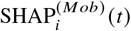,the deep learning model’s predictions. The SHAP values indicate i) greater relative importance of the mobility-weighted sum of neighbours’ cases, compared to the neighbourhood’s own autoregressive process of case generation, at all times where cases are greater than zero, and ii) also that the mobility-weighted sum scales more drastically with the annual epidemics than the autoregressive process (Figure 2E). This suggests a potential greater relative importance of human mobility (versus the autoregressive process) for the persistence of dengue in these urban environments.

Of course, *r*_*i*_ *(t*) and the SHAP mobility influence estimates are not directly comparable, as the SHAP value scales with the magnitude of the data (e.g. Figure 2E), unlike our Bayesian hierarchical model (which models DIRs using population offsets). Therefore, we used the relative SHAP quantity, *r*SHAP_*i*_ *(t*), computed as the ratio between the mobility-weighted influence (to DIR) and autoregressive influence SHAP values (see Materials and methods). We found that there was no one-to-one, linear relationship between *r*_*i*_ *(t*)and *r*SHAP_*i*_ *(t*) (Figure SI 5), perhaps due to the lack of a mechanistic or generative structure in our deep learning model which could constrain the range of plausible values for *r*SHAP_*i*_ *(t*). However, times with greater relative importance of human mobility from the Bayesian model were consistently associated with higher SHAP values for the mobility effect (Figures 3H, SI 5, SI 6), particularly in years with larger dengue outbreaks. Using t-tests, we formally tested this relationship, between *r*_*i*_ *(t*)and *r*SHAP_*i*_ *(t*) across all observations and for each of the 119 neighbourhoods individually (see Figure 3F for Bonferroni-adjusted^[48]^ p-values). For both models, we estimated similar trends for the timings of maximal importance of human mobility (middle stages of season, Figures 3E-F, SI 5, and SI 6).

The maximal relative importance of mobility was most often observed at the midpoint of the annual dengue season (Figure 3E-F), and the relative importance values generally tracked the seasonal epidemic curve (Figure SI 6), yet this estimated pattern varied across seasons (Figure SI 7) and across neighbourhoods (Figures SI 8–SI 9).

Alongside the consistent high values of *r*_*i*_ *(t*) (above 1), these results collectively suggest a potential longer-term trend of human-mobility-driven spread (in urban environments) being consistently more important than internal case generation across all stages of the dengue season, and likely most important in early-to-mid stages of the dengue season, both in more and less severe dengue seasons. Mechanistically, this implies that human-mobility-driven case importations/amplifications may account for sustained circulation of (reported) dengue cases, particularly at seasonal peaks (Figure SI 6).

To further investigate the plausibility of these findings, we explored the human movement patterns using various mixing metrics, such as modularity (from the Louvain and Infomap algorithms) and Shannon’s entropy for the mobility matrix (Figure SI 10). We estimated consistently low modularity (and the Infomap algorithm did not detect any communities) which indicated a well-mixed mobility network in Fortaleza, at least at a neighbourhood level of analysis. Similarly, the entropy was high across all weeks (Median = 7.96, minimum = 7.87, and maximum = 8.06), relative to the maximum achievable of log (119^2^) = 9.56. This high entropy indicates more mixing and more evenly spread flows throughout Fortaleza. Collectively, the results indicate that high mixing may have generated the consistently high importance of human movement and the widespread epidemics.

### 3.3 Space-varying human-mobility-driven influences of neighbourhoods

Different neighbourhoods in a city can play different roles in daily life (e.g. serving as commuter hubs, retail epicentres, or residential areas), and therefore play different roles in seeding and/or sustaining the dengue epidemics of other neighbourhoods. We used our Bayesian hierarchical model, eqs (1)–(9), in rolling windows alongside the derived quantity 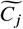,eq. (11), to investigate which neighbourhoods were key contributors (i.e. sources) to the dengue outbreaks in *other* neighbourhoods, and to determine whether these neighbourhoods played consistent source roles within each season and across all seasons. As 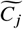 was standardised per window (see Materials and methods), we could compare the neighbourhoods’ (relative) contributions at different stages of dengue seasons. This comparison of neighbourhoods’ contributions over time differs from above, where we were concerned with the estimated time-varying relative importance of human mobility to each neighbourhood’s dengue season, as here we are focused on source-sink dynamics. 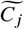 does not represent a neighbourhood’s overall importance for the public health burden in dengue, but rather the human-mobility-driven relative influence of neighbourhood *j* on the epidemic dynamics in all other neighbourhoods.

We found a strong, spatially consistent pattern of high-mobility neighbourhoods consistently being the greatest contributors, in rolling windows, to the epidemics of other neighbourhoods. This pattern was replicated across different stages of individual dengue seasons (Figure 4A), and across all dengue seasons (Figure 4B–C). There was generally little variability in the standardised contributions of individual neighbourhoods across the 258 rolling windows (Figures 4C and SI 11). Indeed, a GAM for median 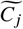 values with a very simple Gaussian linear model structure, neighbourhood-specific random effect, and intercept (i.e. no temporal effects) explained 84% of the variation in 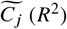.

**Figure 4:**
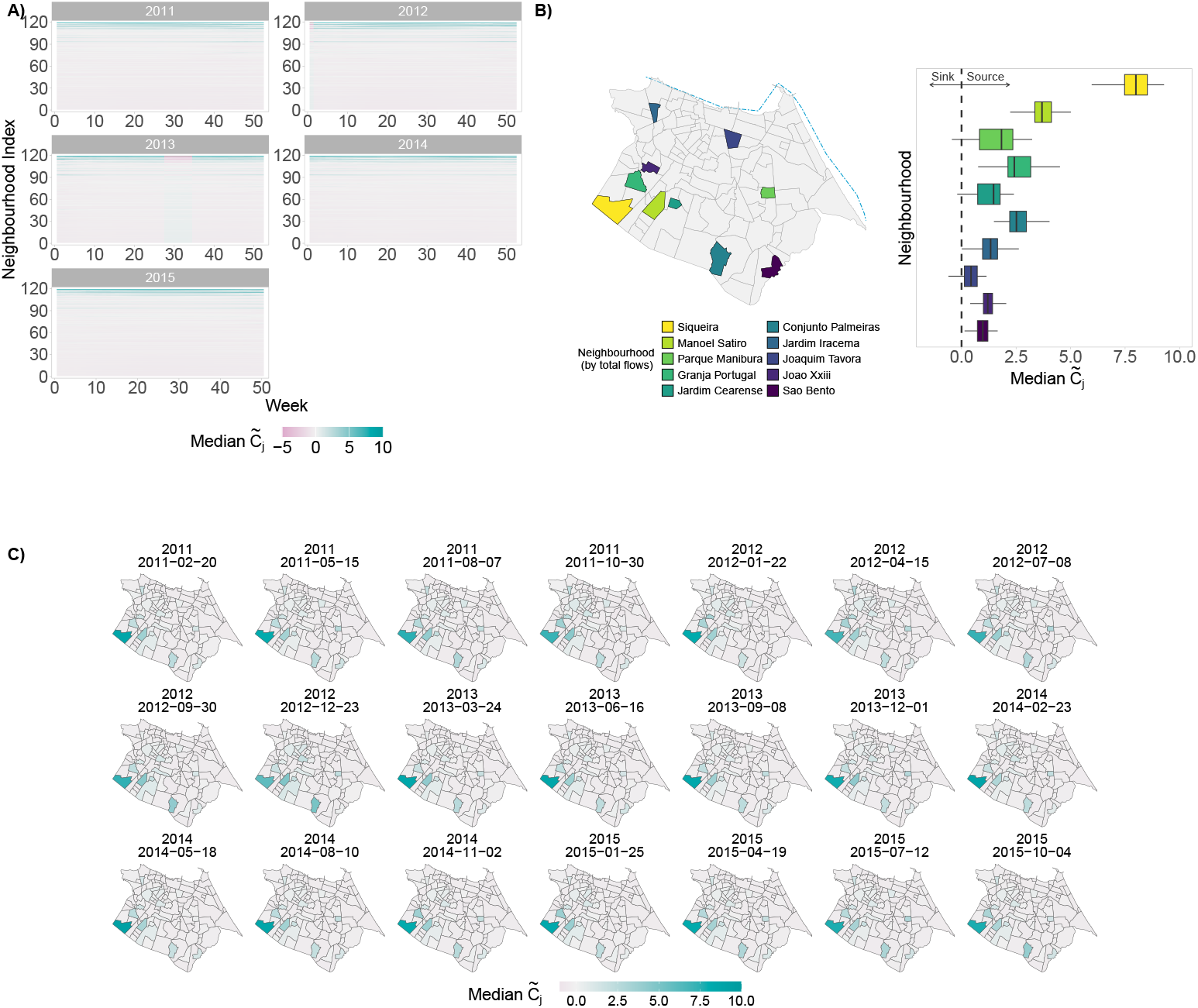
Space-varying source-sink dynamics within and across dengue seasons. Estimated relative (standardised) contributions of individual neighbourhoods to the epidemics of all other neighbourhoods, where estimates are based on the fits of our Bayesian hierarchical model, eqs (1)–(9), and the derived quantity 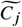,eq. (11). The model was fitted in rolling windows of length 17 weeks to allow for sufficient data for model training, while allowing for potentially time-varying effects. **A)** and **C)** are the median 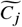 for different neighbourhoods in different rolling windows across 2011–2015, where the week/date visualised represents the maximum date of the rolling window. Neighbourhoods in A) are ordered in ascending order from 1 to 119 by their total annual number of flows (inflows and outflows). Note that in **C)**, we visualised results for only every 12th rolling window (i.e. every 12 weeks) for legibility, and this results in different limits to the legend colours (compared to A). **B)** is a boxplot (right) for the neighbourhoods with the top ten total annual number of flows (left), where for each neighbourhood, all rolling window median 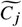 values are summarised. The box and whiskers cover the interquartile range.

Spatially, while intuitive, it is not guaranteed that neighbourhoods with high mobility flows are consistently the greatest contributors. The contributions of generally high-mobility neighbourhoods could be influenced by i) weekly variability in pairwise mobility patterns and/or ii) the stage of the epidemic (as a high-mobility neighbourhood could, in some windows, have relatively few infectious hosts, relative to other neighbourhoods). In our case, both factors, and ii), are apparently not applicable as we found temporally homogeneous mobility patterns (Figure SI 2) and high epidemic synchrony in the city (Figures SI 3 and SI 4). These findings are limited by our dependence on bus transportation data only (see Discussion). For example, the neighbourhood of Siqueira had the greatest inflows (Figure 1B), outflows (Figure 1C), and estimated contributions to other neighbourhoods (Figure 4A–C), and is home to a bus terminal, which could inflate this neighbourhood’s estimated influence on other neighbourhoods.

The only exception to the high relative influence of high-mobility neighbourhoods was during the “peak” of the low-incidence 2013 season, when their estimated relative contribution to other neighbourhoods was lower than lower-mobility neighbourhoods. This phenomenon may be partially explained by the highest cases during this period not being in the highest-mobility neighbourhoods (Figure SI 12) and the lower synchrony of the outbreaks in the low-incidence season of 2013 (Figure SI 4, Median Pairwise Pearson’s Correlation Coefficient^[46]^ *r* = 0.36, Q1 = 0.22, Q3 = 0.49), yet the high-mobility neighbourhoods did not have the lowest cases during this wave (Figure SI 12) and this spatial mobility influence trend was not replicated in the next lower-incidence season of 2014. This phenomenon could, of course, be a modelling limitation as the ELPD LOO indicates poorer generalisability, compared to a model without mobility, during this wave (Figure 2D)

## 4 Discussion

Human movement is widely understood as an important driver of many respiratory, vector-borne, waterborne, sexually transmitted and other infectious diseases^[53,54,55]^, shaping the propagation of infectious diseases across space and time. However, for dengue epidemics in large cities, it was less clear how i) the relative importance of human mobility and the human-mobility-driven source-sink dynamics of different neighbourhoods, vary within and across individual dengue seasons. Here, we provided multi-model methods to investigate these questions, illustrating our application in high spatial resolution for urban dengue outbreaks in Fortaleza, Brazil.

Understanding *when* human mobility is most important in an outbreak has practical consequences for public health policy. This can inform the optimal timing for public health interventions (e.g. vector control strategies) for endemic, epidemic, and pandemic pathogens. Understanding *where* human movement is seeding, expanding and/or sustaining outbreaks of other geographies can enable more targeted and cost-effective interventions. For example, in our study, while we estimated that human mobility was consistently important across neighbourhoods throughout a dengue season, we identified a high relative importance i) of human mobility at the middle stages and peaks of dengue seasons and ii) of particular neighbourhoods to geographic spread in each dengue season. Targeting vector control interventions to reduce human-vector contact within such times and neighbourhoods could have an outsized, positive impact on bringing the epidemic under control.

Beyond public health interventions, such insights could also be used to inform design of human movement data collection strategies. Routine mobility data collection and sharing, particularly in low- and middle-income countries (LMICs), is challenging^[21]^ (due to economic, political, and logistical barriers) and often infeasible, therefore suggesting that more cost-effective and targeted data collection is needed. Although our focus here has not been on forecasting (see^[18,40,56,57,58]^) nor invasion dynamics (see^[20,59,60]^), we have demonstrated consistently strong (out-of-sample) predictive ability of our models via the enhancement of spatiotemporal predictions of dengue incidence by using human mobility data. Therefore, incorporating the spatial structure induced by human mobility in invasion and forecasting models is likely important for more accurate forecasts that can be used for short-term outbreak response and longer-term seasonal planning.

Our results complement previous work using similar and other data streams to investigate other questions about the influences of human mobility on shaping the spread of infectious diseases. In addition to public transportation data, researchers have used human movement data from air and water travel^[61]^, population censuses^[62]^, and mobile phones^[63,64]^ to investigate questions of disease spread on subnational, national, and global scales. Future work may look at understanding how best to integrate these heterogeneous mobility data sources with our methods and analyses (potentially extending research questions to roles of long-distance human movements), as well as the net information gains (including corresponding costs and potential data biases) from such additional data collection. These net information gains will depend on the setting-specific research questions (e.g. early warning detection, nowcasting, or forecasting) and the time and region, yet quantifying the information gains is crucial for encouraging and guiding data collection. This could be a stepping stone towards addressing our aforementioned barriers caused by the lack of subnational human mobility data, particularly in resource-limited LMICs^[21,65]^. Understanding data information gains is also important as different model-based proxies^[21]^ and mobility data streams^[65]^ can induce biases in the modelling estimates that are ultimately used to inform public health authorities, although there are recent advances in the accuracy of mobility data proxies (e.g. gravity-like models^[66]^) and integrating heterogeneous mobility data streams^[67]^.

The consistently high relative importance of human mobility (and peaks coinciding with seasonal epidemic peaks) and of specific source neighbourhoods makes intuitive sense within the context of a large, highly urbanised, well-mixed, and commuter-centric city such as Fortaleza. Mechanistically, once the seasonal urban dengue outbreak becomes established, the movement of humans and thus, provision of infectious hosts, apparently allows for the epidemic surges (i.e. expansion) and sustained spread across neighbourhoods. In previous work, arbovirus risk (for dengue, Zika, and chikungunya) in Fortaleza (albeit focused on 2015 onwards) had estimated spatial clusters of cases^[68]^, including for clusters of neighbourhoods over longer distances (up to 6.8 km). The authors hypothesised that human mobility likely allowed infected individuals to regularly import viruses to distant areas in Fortaleza. Similarly, infestation surveys, which estimate *Ae. aegypti* density, frequently indicate a high level of vector adaptation across Fortaleza^[24]^. Interpreted alongside a consistent role for human movement, this suggests that, even in less suitable climatic conditions (e.g. dry season), the mosquito vector could be present near homogeneously across space and time. This example underlines the importance of understanding the history of dengue transmission when applying our framework and interpreting results in other settings. In recently affected cities (e.g. Buenos Aires, Argentina), the likely fluctuations in mosquito vector density (as the effects of climate change materialise^[60,69]^) could modulate mobility effects and yield very different influences of human movement on urban dengue epidemic dynamics.

Meanwhile, for COVID-19 Aguilar et al. ^[70]^ compared hierarchical cities with sprawled/well-mixed cities, and found that epidemic spread was less rapid in sprawled cities. Interpreting our results, albeit for a different infectious disease, alongside these findings, we suspect that the high mixing across the city (Figure SI 10) may have supported the relatively long dengue seasons frequently observed in Fortaleza (Figure SI 3B). This may explain why we estimated a high relative importance of mobility in multiple individual dengue seasons. Further work could investigate whether these results generalise in other environments – different mobility patterns, different spatial scales of cities, and different socio-environmental settings.

Our findings align with research that documented consistently superior overall predictive performance of mobility-informed models in Fortaleza^[18,19]^, other cities (e.g. 12 cities in Guangzhou province, China^[13]^), and at national levels (e.g. in Pakistan^[12]^ and Vietnam^[8]^). Our research also builds on such research by disentangling the relative importance and influences of human movement for different stages of the epidemic and for different neighbourhoods. and *r*SHAP (*t*), eq. (13)) and of different neighbourhoods (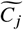,eq. (11)) could be useful. These could be paired For future studies, our unitless measures of the time-varying relative importance of human mobility (*r*_*i*_ (*t*), eq. (10), with new formulae for other unitless measures of epidemic properties such as time-varying reproduction numbers *R (t*) (e.g. for dengue^[71,72]^) to further quantify how the relative importance of human mobility and the influence of different neighbourhoods evolve with varying epidemic strength (as measured by *R (t*), the number of new infections generated by a typical individual, see^[73]^).

The models that we develop are new for modelling the time- and space-varying roles and effects of human movement on dengue epidemics, yet the models remain conceptually simple. The key advance is the ability to derive mathematical quantities (together with robust uncertainties) for the importance and influence of human mobility from a generative, Bayesian model, and to blend the results for these quantities with those using our derived formulae for game-theoretic quantities (i.e. SHAP and rSHAP values). This multi-model procedure for extracting spatiotemporal insights (by testing theories and synthesising evidence across disciplines) enhances robustness of causal interpretations^[44]^, and our conceptual advances are generalisable to other settings (e.g. other infectious diseases) and other research questions of relative importance (e.g. climatic versus socioenvironmental influences).

Our analysis has several data limitations and there are many future avenues of research. First, we used weekly bus transportation data from 2015 to represent movements for all years. Although there is potential for representation bias from any form of mobility data^[65]^ and we advocate for greater availability of more regular data, previous analyses have shown that these data are representative for a wide range of explanatory and predictive research questions (e.g. crime rates, dengue epidemics and police allocation)^[18,27,28,29]^. Similarly, there were no substantial time variations in flows throughout the weeks of 2015 (Figure SI 2) and our mobility-informed models consistently enhanced performance across information and predictive performance criteria, hinting at the generalisability of these data (beyond 2015) and at the potential value of intermittent collection and sharing of such data. However, the aggregated bus data only capture shorter-distance movements (as opposed to importations from inward travel to the city) are not linked to demographic or social identifiers, nor specific disease status, and future work could explore the additional value or differences in results from integrating other individual-level human movement indicators (e.g. mobile phone data) and long-distance movement indicators (e.g. flight data). Such data sources may also have greater variability throughout the dengue season and provide valuable additional information for understanding the dynamics of invasion and geographical spread. Our models did not explicitly account for within-neighbourhood mobility, as bus transportation data were more suitable (within-neighbourhood bus trips were rare) for our studied research question – comparing imported cases with those generated within neighbourhoods, which means that within-neighbourhood cases can therefore account for human, biological, and environmental effects. Future work, likely relying on shorter-range movement data (compared to bus transportation data), could explore the relative importance of within-neighbourhood mobility over space and time. This would be instructive for better understanding of local community transmission in large informal settlements, which could be supported by the movement of infectious individuals on foot or motorcycles, coupled with the spread of mosquito breeding sites.

Further data limitations include our usage of observed dengue case data which are an imperfect measure of true dengue infections. The case data may be sensitive to reporting delays and/or underreporting in time or space (e.g. due to health centre availability or reporting capacity/propensity), which we attempted to proxy by using neighbourhood-specific random effects. These issues may also be exacerbated by the high spatial resolution of our study, yet this is balanced against the benefits of i) avoiding potential spatial aggregation artefacts/effects and ii) providing high-resolution insights for targeted policy questions.

Meanwhile, we did not have access to serotype or seroprevalence data. This may limit the robustness of our illustrative application as all four DENV serotypes have circulated in Fortaleza and 2011 and 2012 outbreaks were caused by different serotypes (the reintroduction and introduction of DENV-1 and DENV-4 respectively), which was the first time that a major Brazilian city experienced a serotype switch in two consecutive annual dengue epidemics. In 2015, DENV-1 was dominant once again. Overall, our results are likely not confounded by more recent introductions of Zika, and chikungunya viruses (which would induce misclassifications, yet these were almost simultaneously first reported in 2015)^[68]^. We also used only the one-week-lagged cases and did not model the mosquito population directly, and therefore we only approximated the true causal dynamical processes (e.g. biological mechanisms) of DENV transmission. Further causal limitations include the limitations of explainability metrics (such as SHAP values) for deep learning models when disentangling causal effects^[42,45]^ (see Materials and methods). In future work, contingent on the availability of new data streams (seroprevalence, genomic, higher spatial resolution climate, or mosquito catch data), researchers could consider alternative networked mechanistic (or biomathematical) and/or deep learning models. These models could enhance the causal realism with explicit time- and space-varying properties (e.g. generation time and serial interval distributions^[71,72]^). Similarly, models could incorporate additional human influences (e.g. population immunity^[10]^, access to clean drinking water^[7,74]^ and circulating serotypes^[8,11]^), climatic drivers (e.g. interactions with precipitation, drought, humidity, or temperature^[7,75]^) and/or mosquito factors (e.g. mosquito density^[6,76]^).

In conclusion, our theoretical advances (evidence synthesis across multiple models) and findings (influences of human movement) can be a foundation for i) future studies that improve our understanding of the complex processes of dengue epidemics in different settings and ii) future public health policy that targets limited resources for dengue epidemic control.

## Data Availability

All code and data used in this analyses are available at https://github.com/cathalmills/mobility_relative_importance

http://dx.doi.org/10.1098/rsif.2020.0691

