## Supplementary material for "The time- and space-varying roles of human mobility in shaping urban dengue epidemics"

### A Appendix

#### A.1 Notation and abbreviations

Table SI 1: **Model abbreviations and description.**

| Abbreviation | Definition |
|---|---|
| Bayesian | A Bayesian hierarchical model for dengue cases with mobility effects and neighbourhood-specific autoregressive processes for lagged cases. |
| No-Mob-Bayesian | A Bayesian hierarchical model for dengue cases with neighbourhood-specific autoregressive processes and no mobility effects for lagged cases. |
| DL | A foundational time series model (TimeGPT[36]) with mobility effects and autoregressive processes for lagged cases. |
| No-Mob-DL | A foundational time series model (TimeGPT[36]) with autoregressive processes for lagged cases. |
| Baseline | A baseline forecasting model for comparison and model validation (see[41,77]) . |

Table SI 2: **List of abbreviations and notations.**

| Acronym | Description |
|---|---|
| AR(1) | Autoregressive process of order one |
| CRPS | Continuous Ranked Probability Score |
| DENV | Dengue Virus |
| DIR | Dengue Incidence Rate |
| DL | Deep Learning |
| ELPD-LOO | Expected Log (pointwise) Predictive Density using Leave-One-Out (LOO) cross-validation |
| GAM | Generalised Additive Model |
| GCV score | Generalised Cross-Validation score |
| IBGE | Brazilian Institute of Geography and Statistics |
| LOO | Leave-One-Out cross-validation |
| MCMC | Markov Chain Monte Carlo |
| ML | Machine Learning |
| NegBin | Negative Binomial Distribution |
| $r_i(t)$ | Ratio of mobility-generated contributions to a neighbourhood versus a neighbourhood's autoregressive process |
| $r\text{SHAP}_i(t)$ | Our relative SHAP value for the estimated importance of human mobility effects |
| rWIS | Relative Weighted Interval Score |
| SHAP | SHapley Additive exPlanations (used for model interpretability) |
| SIMDA | Sistema de Monitoramento Diário de Agravos (Daily Disease Occurrence Monitoring System) |
| WAIC | Widely Applicable Information Criterion |
| WIS | Weighted Interval Score |

#### A.2 Alternative candidate statistical models

We now consider alternative Bayesian hierarchical approaches to modelling $y_{it}$, the reported neighbourhood-level ($i$) weekly ($t$) case counts of dengue.

**Statistical model I: Space-varying mobility effects**

Our first framework included a mobility effect for each focal neighbourhood $i$, and weighted the past DIR per 100,000 of all neighbourhoods ($\eta_{j,t-1}$), including the focal neighbourhood itself, by the proportion of total flows originating from neighbourhood $j$ to the focal neighbourhood $i$:

$$
\begin{aligned}
y_{it} &\sim NegBin(\mu_{it}, \phi), \\
\log(\mu_{it}) &= \log(P_{it}) + \log(\eta_{it}), \\
\log(\eta_{it}) &= \alpha + \left( \beta_{\text{Mobility}[i]} \cdot \sum_{j \in \mathcal{N}(i) \cup i} \frac{\mathrm{f}_{t-1}^{(j,i)}}{\sum_{h \in \mathcal{N}(i) \cup i} \mathrm{f}_{t-1}^{(h,i)}} \cdot \eta_{j,t-1} \right) + \beta_{\text{Year}[t]} + \beta_{\text{Nbr}[i]}, \\
\beta_{\text{Mobility}[i]} &\sim N(\beta_{\text{Mobility}}, \sigma_{\text{Mobility}}), \\
\beta_{\text{Mobility}} &\sim N(0,1), \\
\sigma_{\text{Mobility}} &\sim \text{Half-Normal}(0,1), \\
\alpha &\sim N(0,1), \\
\beta_{\text{Year}} &\sim N(0,1), \\
\beta_{\text{Nbr}} &\sim N(0,1), \\
\phi^{-1} &\sim \text{Half-Normal}(0,1),
\end{aligned}
\tag{14}
$$

where $\mu_{it}$ denotes the mean parameter of the Negative Binomial distribution and $\phi$ is the reciprocal overdispersion parameter. We used a population offset $\log(P_{it})$ such that $\eta_{it}$ represented the DIR per 100,000 population.

The DIR was then modelled with i) an intercept $\alpha$, ii) a neighbourhood-specific mobility fixed effect controlled by $\beta_{\text{Mobility}[i]}$, the lagged DIR of all neighbourhoods ($\mathcal{N}(i) \cup i$ including the focal neighbourhood $i$), weighted by the inter- and intra-flows, iii) a year-to-year random effect (i.e. $\beta_{\text{Year}}$ is a varying intercept for each year), and iv) a neighbourhood-specific random effect (i.e. $\beta_{\text{Nbr}}$ is a varying intercept for each neighbourhood). So, $\beta_{\text{Mobility}[i]}$ is the neighbourhood-specific effect of past DIR across all neighbourhoods, weighted by mobility flows. Then, the common mean and standard deviation of these neighbourhood-specific effects are hyperparameters ($\beta_{\text{Mobility}}$ and $\sigma_{\text{Mobility}}$ respectively) which are given hyperprior distributions. All Normal prior distributions are presented in mean-standard deviation representation.

**Statistical model II: Space-varying mobility with autoregressive processes**

Our second model differs to that specified in eq. (14), as it allowed us to isolate the roles played by the autoregressive process of outbreaks within neighbourhoods and the mobility flows of (reported) infected individuals from other neighbourhoods.

$$
\begin{aligned}
y_{it} &\sim NegBin(\mu_{it}, \phi), \\
\log(\mu_{it}) &= \log(P_{it}) + \log(\eta_{it}), \\
\log(\eta_{it}) &= \alpha + \left( \beta_{\text{Mobility-Nbr[i]}} \cdot \sum_{j \in \mathcal{N}(i)} \frac{\mathrm{f}_{t-1}^{(j,i)}}{\sum\limits_{h \in \mathcal{N}(i)} \mathrm{f}_{t-1}^{(h,i)}} \cdot \eta_{j,t-1} \right) + (\beta_{\text{AR[i]}} \cdot \eta_{i,t-1}) + \beta_{\text{Year}[t]} + \beta_{\text{Nbr}[i]}, \\
\beta_{\text{Mobility-Nbr}[i]} &\sim N(\beta_{\text{Mobility-Nbr}}, \sigma_{\text{Mobility-Nbr}}), \\
\beta_{\text{Mobility-Nbr}} &\sim N(0,1), \\
\sigma_{\text{Mobility-Nbr}} &\sim \text{Half-Normal}(0,1), \\
\beta_{\text{AR}[i]} &\sim N(\beta_{\text{AR}}, \sigma_{\text{AR}}), \\
\beta_{\text{AR}} &\sim N(0,1), \\
\sigma_{\text{AR}} &\sim \text{Half-Normal}(0,1), \\
\alpha &\sim N(0,1), \\
\beta_{\text{Year}} &\sim N(0,1), \\
\beta_{\text{Nbr}} &\sim N(0,1), \\
\phi^{-1} &\sim \text{Half-Normal}(0,1)
\end{aligned}
\tag{15}
$$

where $\mu_{it}$, $\phi$, $P_{it}$, $\eta_{it}$, $\alpha$, $\beta_{\text{Year}}$, and $\beta_{\text{Nbr}}$ are all defined as above. $\beta_{\text{Mobility-Nbr}[i]}$ is a neighbourhood-specific parameter controlling the effect of the weighted and lagged DIR of other neighbourhoods (excluding the focal neighbourhood), weighted by the inter-neighbourhood mobility flows. $\beta_{\text{AR[i]}}$ is the neighbourhood-specific parameter controlling a (potentially non-stationary) autoregressive process of order one, AR(1), for each neighbourhood $i$. As above, a hierarchical structure was imposed such that the mobility effect was modelled with partial pooling (common mean of $\beta_{\text{Mobility-Nbr}}$ and standard deviation of $\sigma_{\text{Mobility-Nbr}}$), and an identical procedure was performed for the autoregressive process (common mean of $\beta_{\text{AR}}$ and standard deviation of $\sigma_{\text{AR}}$).

**Statistical model III: Time-varying mobility with neighbourhood autoregressive effects**

Let $y_{it}$ be the reported neighbourhood-level ($i$) weekly ($t$) case counts of dengue which was modelled with a Negative Binomial (NegBin) likelihood as follows:

$$
\begin{aligned}
y_{it} &\sim NegBin(\mu_{it}, \phi), \\
\log(\mu_{it}) &= \log(P_{it}) + \log(\eta_{it}), \\
\log(\eta_{it}) &= \alpha + \left( \beta_{\text{Mobility-Nbr}[t]} \cdot \sum_{j \in \mathcal{N}(i)} \frac{\mathrm{f}_{t-1}^{(j,i)}}{\sum\limits_{h \in \mathcal{N}(i)} \mathrm{f}_{t-1}^{(h,i)}} \cdot \eta_{j,t-1} \right) + \beta_{\text{AR}[i]} \cdot \eta_{i,t-1} + \beta_{\text{Year}[t]} + \beta_{\text{Nbr}[i]}, \\
\beta_{\text{Mobility-Nbr}[1]} &\sim N(0,1), \\
\beta_{\text{Mobility-Nbr}[t]} &\sim N(\beta_{\text{Mobility-Nbr}[t-1]}, \sigma_{\text{Mobility-Nbr}}), t \geq 2, \\
\sigma_{\text{Mobility-Nbr}} &\sim \text{Half-Normal}(0,1), \\
\beta_{\text{AR}[i]} &\sim N(\beta_{\text{AR}}, \sigma_{\text{AR}}), \\
\beta_{\text{AR}} &\sim N(0,1), \\
\sigma_{\text{AR}} &\sim \text{Half-Normal}(0,1), \\
\alpha &\sim N(0,1), \\
\boldsymbol{\beta}_{\text{Year}} &\sim N(0,1), \\
\boldsymbol{\beta}_{\text{Nbr}} &\sim N(0,1), \\
\phi^{-1} &\sim \text{Half-Normal}(0,1),
\end{aligned}
\tag{16}
$$

where $\mu_{it}$, $\phi$, $P_{it}$, $\eta_{it}$, $\alpha$, $\beta_{\text{Year}}$, $\beta_{\text{Nbr}}$, $\beta_{\text{AR}[i]}$, $\beta_{\text{AR}}$ and $\sigma_{\text{AR}}$ are all defined as above. Here, we modelled the effect of mobility to be spatially constant yet temporally evolving by allowing $\beta_{\text{Mobility-Nbr}}$ to follow a Gaussian Random Walk with standard deviation $\sigma_{\text{Mobility-Nbr}}$. Each neighbourhood was again given its own autoregressive process and varying intercept.

#### A.3 Results

##### A.3.1 Plots of data

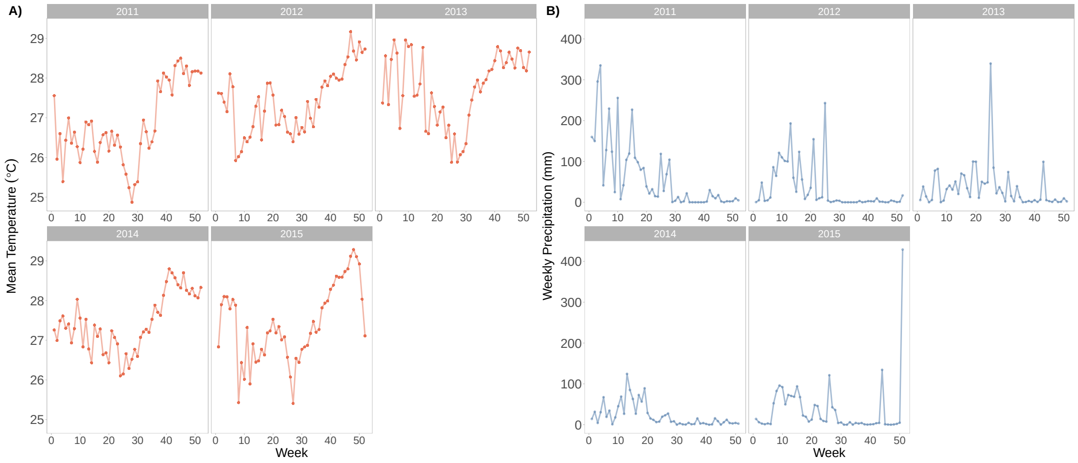


Figure SI 1: **Weekly mean temperature and total precipitation in Fortaleza, 2011–2015: A)** Weekly mean temperature and **B)** weekly total precipitation for the city of Fortaleza, as per the ERA-5 dataset [30], where the variables were calculated as the average across the area within the Fortaleza city boundaries.

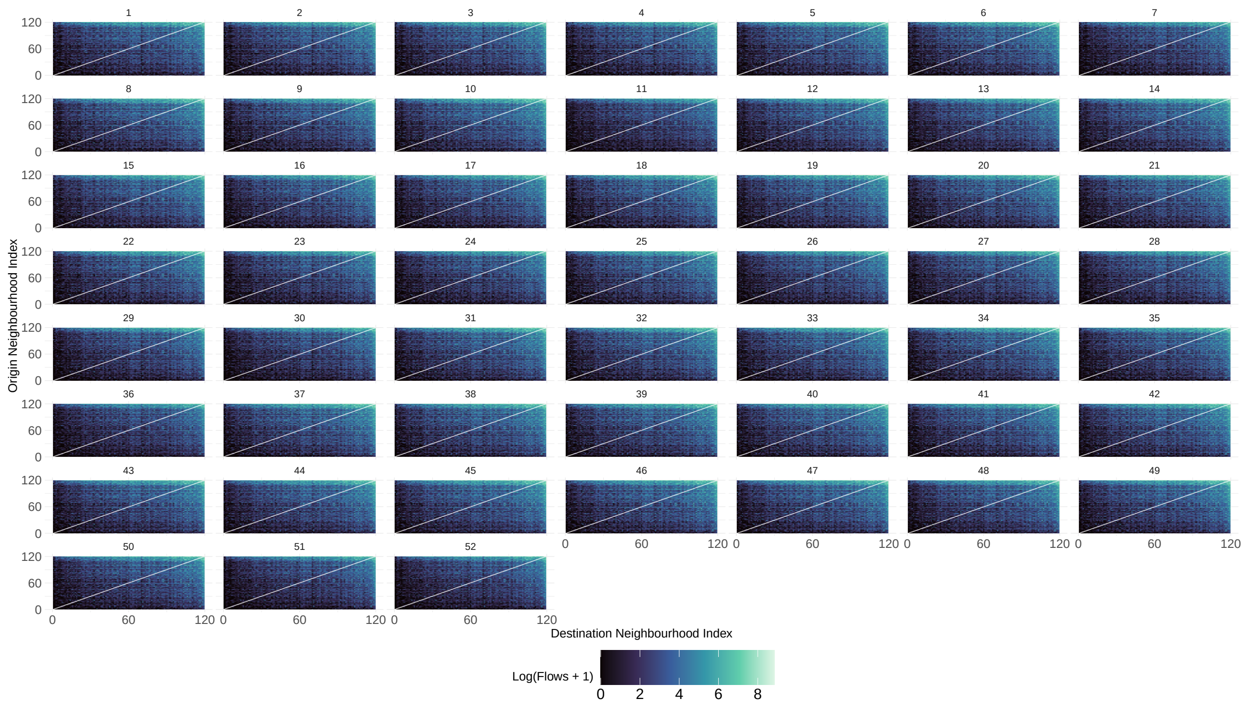


Figure SI 2: **Weekly human movement in Fortaleza, Brazil:** The number of inflows and outflows between two neighbourhoods per week (facets) in 2015, where the neighbourhood index (for both axes) is consistent with other plots (for comparison purposes); ordered in ascending order by their total flows (inflows and outflows).

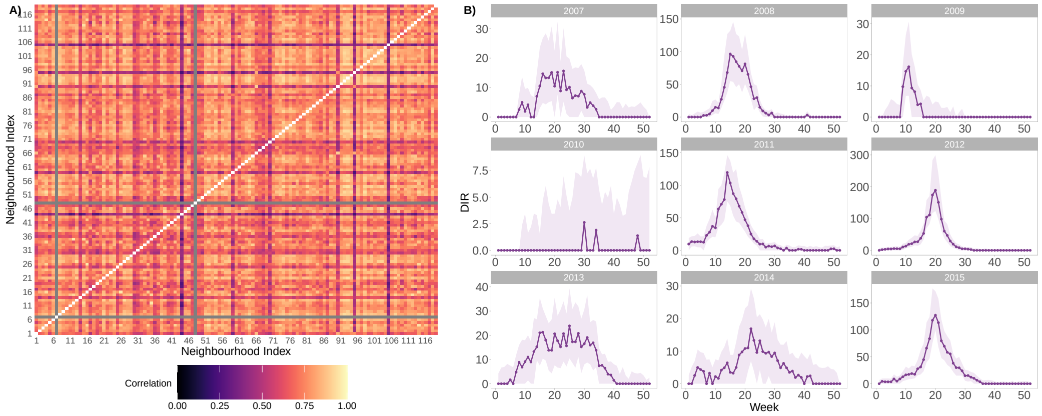


Figure SI 3: **Epidemic synchrony and dengue cases from 2007–2015:** **A)** The Pearson's correlation coefficients [46] between the incident time series (using DIR for comparability) of each pair of the 119 neighbourhoods, where neighbourhoods are ordered in ascending order by their total flows (inflows and outflows). **B)** The median DIR (solid line) is visualised alongside upper and lower quartiles (ribbon) of DIR values for each time point across 2007–2015.

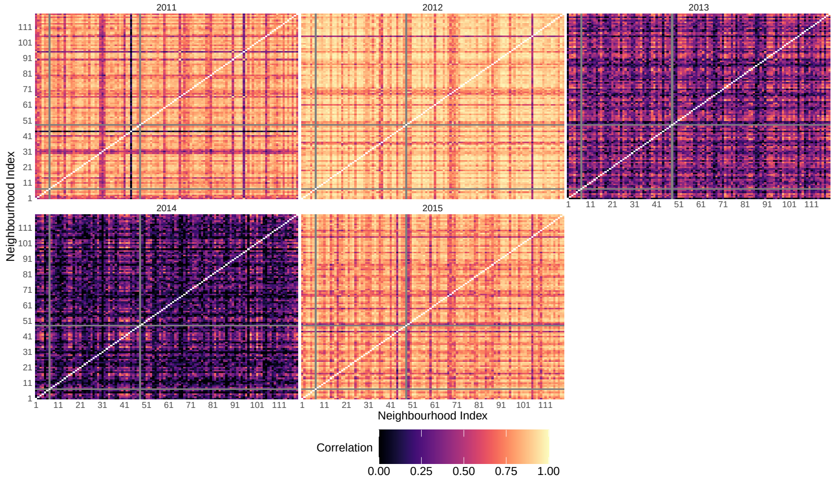


Figure SI 4: **Within-year epidemic synchrony from 2007–2015:** The Pearson's correlation coefficient [46] between the incident time series (using DIR for comparability), per year, of each pair of the 119 neighbourhoods, where neighbourhoods are ordered in ascending order by their total flows (inflows and outflows). The epidemic synchrony for the full studied period is shown in Figure SI 3A).

##### A.3.2 Time-varying relative importance of human mobility

The following section contains results which complement Section 3.2.

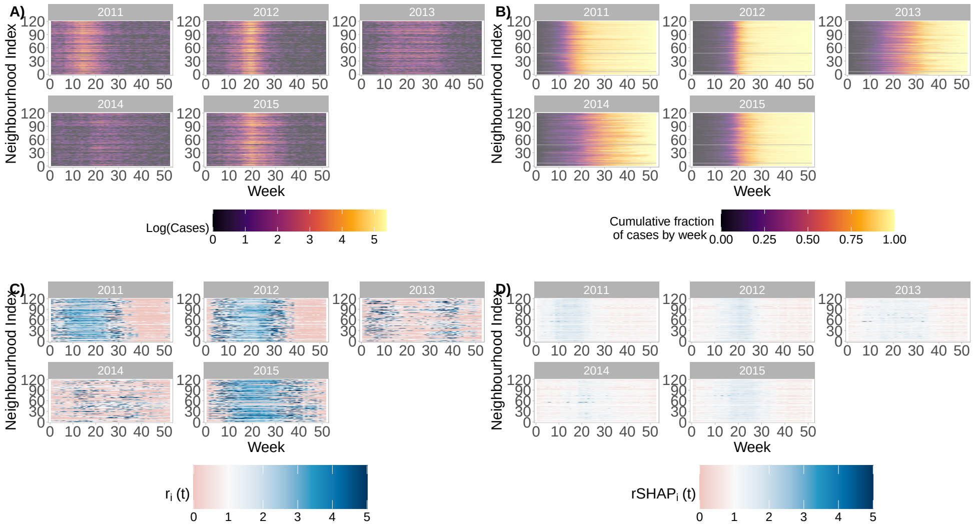


Figure SI 5: **Time-varying relative importance of human mobility by year:** Across the testing period and for each of the 119 neighbourhoods, we visualise the estimated relative importance of human-movement-based importations of dengue cases, versus the neighbourhoods' own autoregressive/internal generation of new dengue cases. **A)** and **B)** visualise the observed cases (on a logarithmic scale) and cumulative fraction of annual cases observed for individual weeks across 2011-2015. Neighbourhoods are ordered in ascending order by their total annual number of flows (inflows and outflows). **C** and **D)** show the estimated relative importance from our Bayesian hierarchical and deep learning models respectively, where values of $r_i(t)$ and $r\text{SHAP}_i(t)$ greater than one indicate greater relative importance of the case importations (see Materials and methods). Figure 3C-D is the analogous continuous version of these estimates of $r_i(t)$ and $r\text{SHAP}_i(t)$.

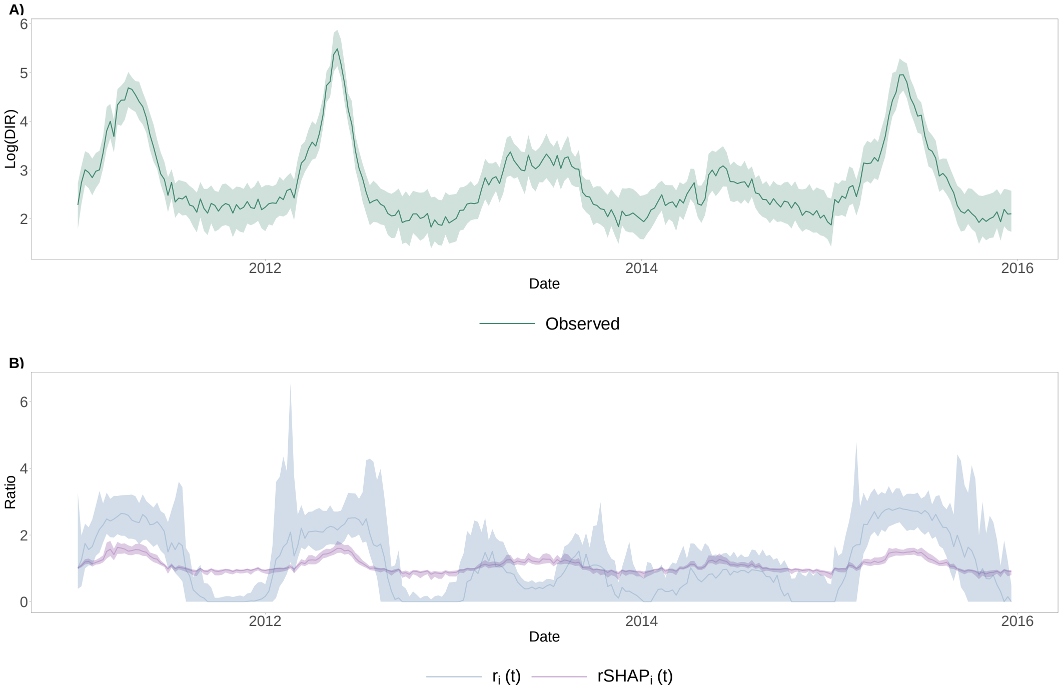


Figure SI 6: **Time-varying relative importance of human mobility by different models, alongside observed case data: A)** Median dengue incidence rate (DIR) on a logarithmic scale is visualised alongside **B)** $r_i(t)$ and $r\text{SHAP}_i(t)$, the model-based estimates for the time-varying relative importance of human mobility.

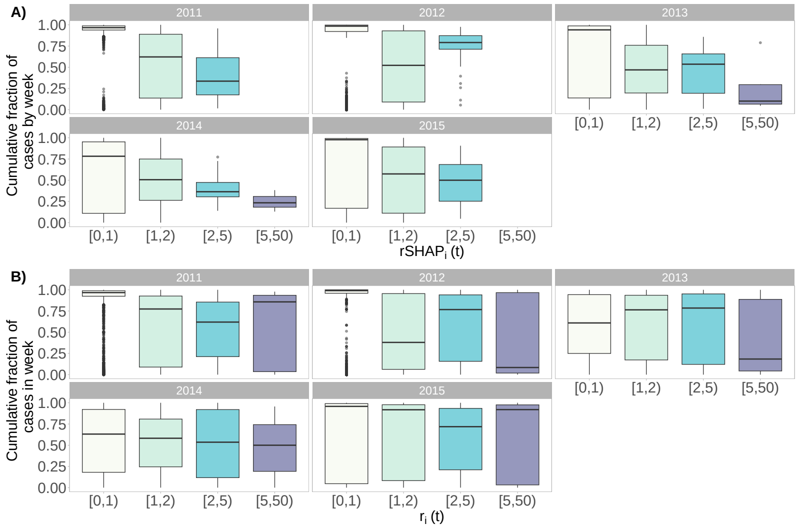


Figure SI 7: **Time-varying importance of human mobility by epidemic stage in different years:** For each year (2007–2015), a summary of the cumulative fraction of annual cases observed by a given week is visualised against the **A)** corresponding deep learning model's estimates and **B)** the Bayesian hierarchical model's estimates for the relative importance of human mobility for each neighbourhood's epidemic dynamics.

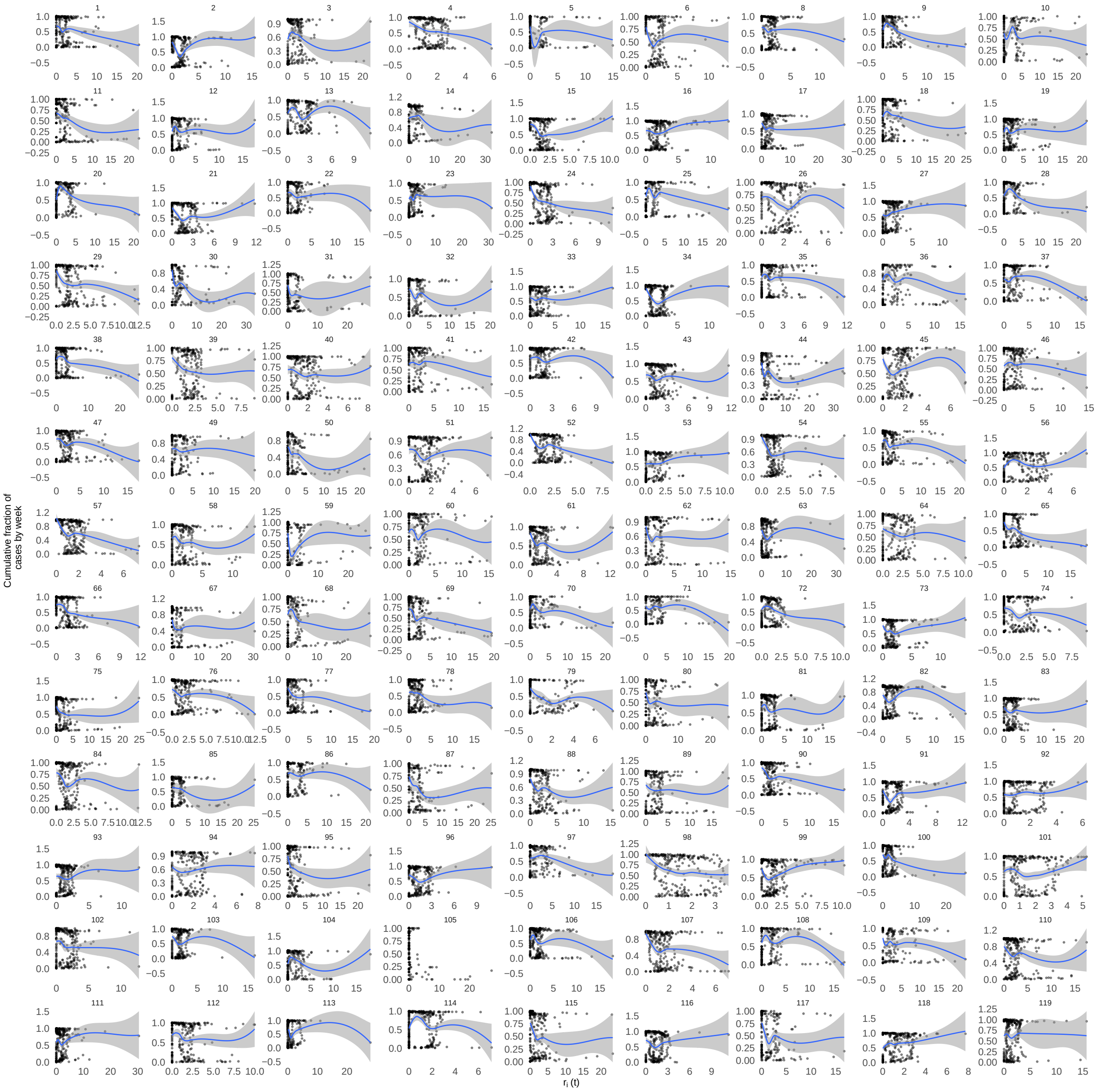


Figure SI 8: **Bayesian model's estimated time-varying importance of human mobility, by epidemic stage in different neighbourhoods:** For each neighbourhood (indexed 1 to 119, in ascending order by mobility flows), the cumulative fraction of annual cases observed by a given week is visualised against the corresponding Bayesian hierarchical model's estimates for the relative importance of human mobility for the neighbourhood's epidemic dynamics. The blue line represents the conditional mean and the shaded band represents the 95% confidence interval (calculated using bootstrap resampling) for each $r_i(t)$, calculated using loess (Local Polynomial Regression Fitting).

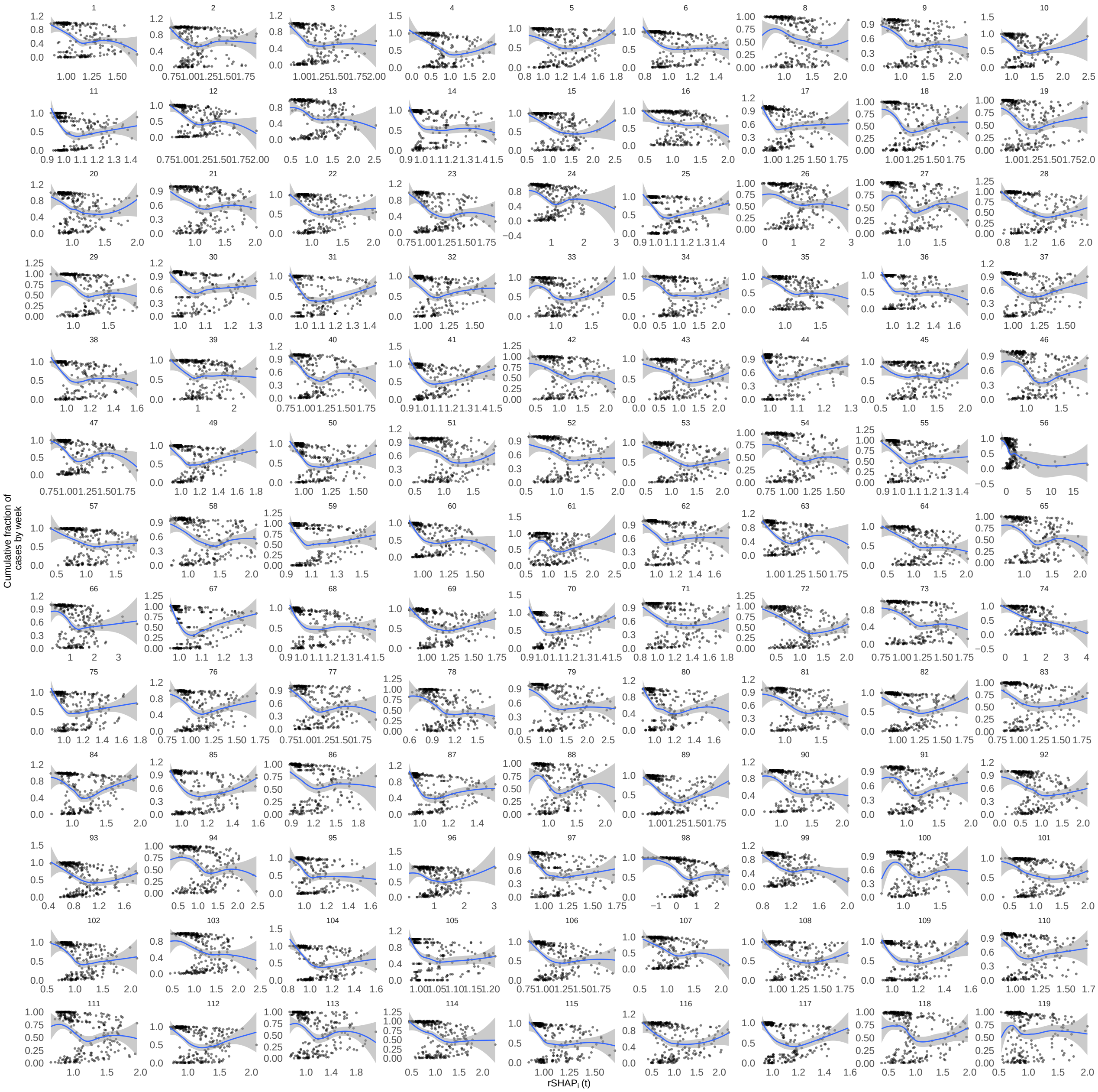


Figure SI 9: **Deep learning model's estimated time-varying importance of human mobility, $r\mathbf{SHAP}_i(t)$ by epidemic stage in different neighbourhoods:** For each neighbourhood (indexed 1 to 119, in ascending order by mobility flows), the cumulative fraction of annual cases observed by a given week is visualised against the corresponding Bayesian hierarchical model's estimates for the relative importance of human mobility for the neighbourhood's epidemic dynamics. The blue line represents the conditional mean and the shaded band represents the 95% confidence interval (calculated using bootstrap resampling) for each $r\mathrm{SHAP}_i(t)$, calculated using loess (Local Polynomial Regression Fitting).

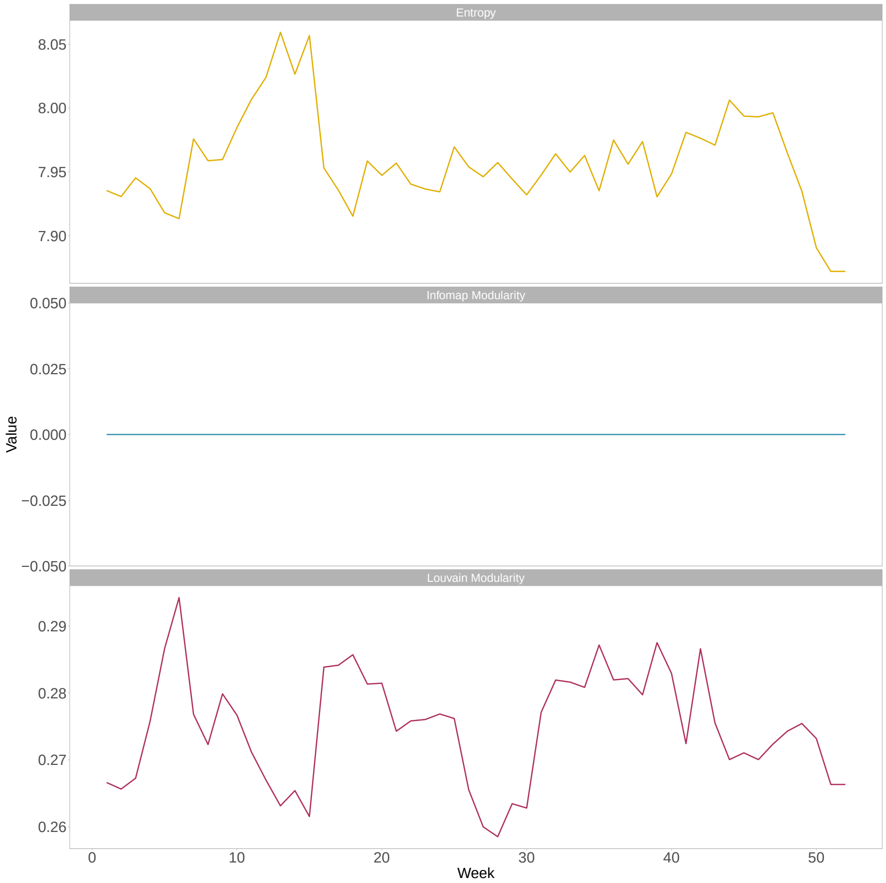


Figure SI 10: **Mixing patterns in Fortaleza by different algorithms:** We quantified mobility clustering using entropy, Infomap modularity, and Louvain modularity, where higher values of entropy and lower values of modularity indicate greater mixing and less mobility clustering. The entropy-based results are constrained by an upper bound of $\log(119^2) = 9.56$, the Infomap modularity indicates no clusters identified, and Louvain modularity ranges from -1 to 1, with values close to zero indicating low clustering.

##### A.3.3 Space-varying human-mobility-driven roles of neighbourhoods

The following section contains results which complement Section 3.3.

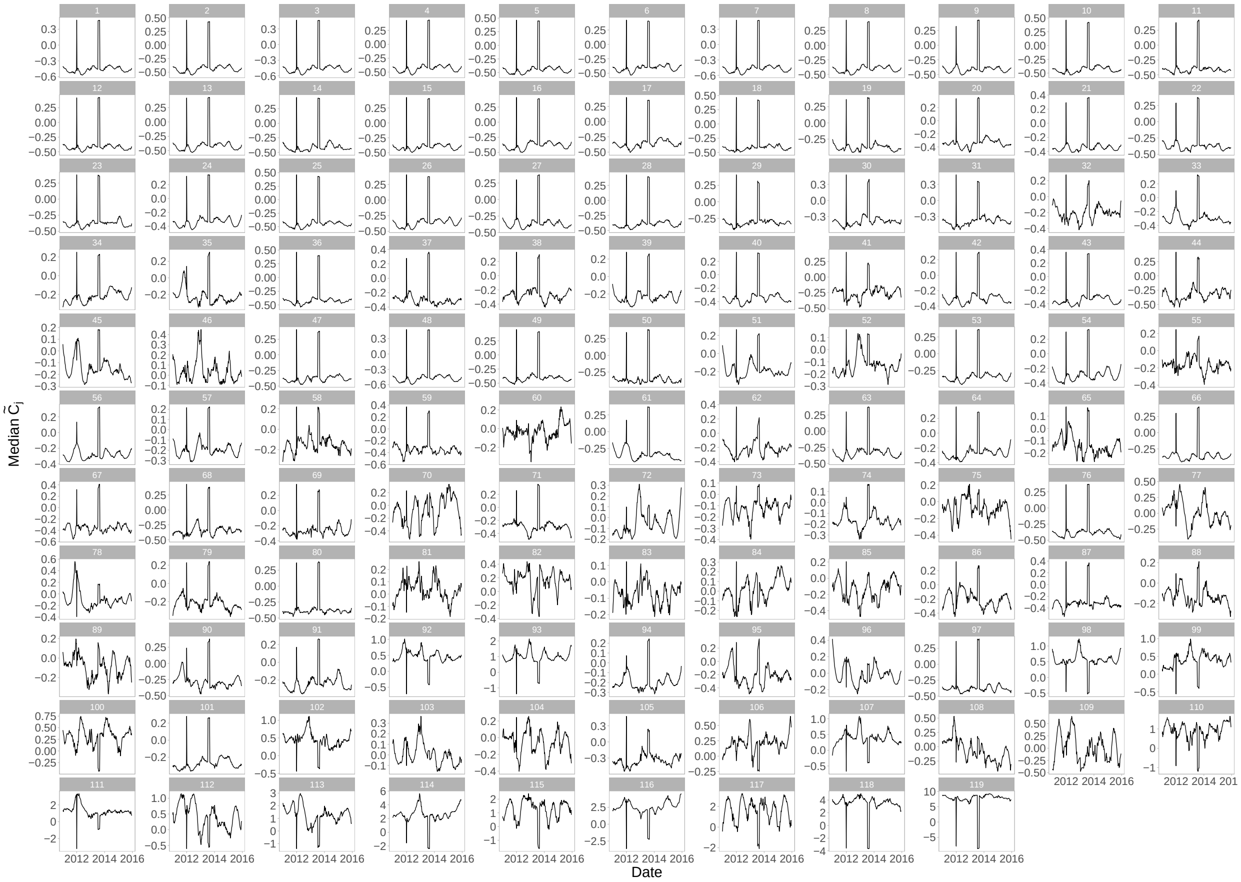


Figure SI 11: **Estimated time-varying contributions of neighbourhoods:** For each neighbourhood $j$, $\widetilde{C_j}$, is shown where the date on the x-axis represents the maximum date for that 17-week trailing window. $\widetilde{C_j}$ is the Bayesian model's estimate for the standardised contribution of neighbourhoods $j$ to the dengue epidemics of all other neighbourhoods. This can be compared across different rolling windows due to the standardisation. Note the different y-axis to preserve the different magnitudes of contributions from different neighbourhoods.

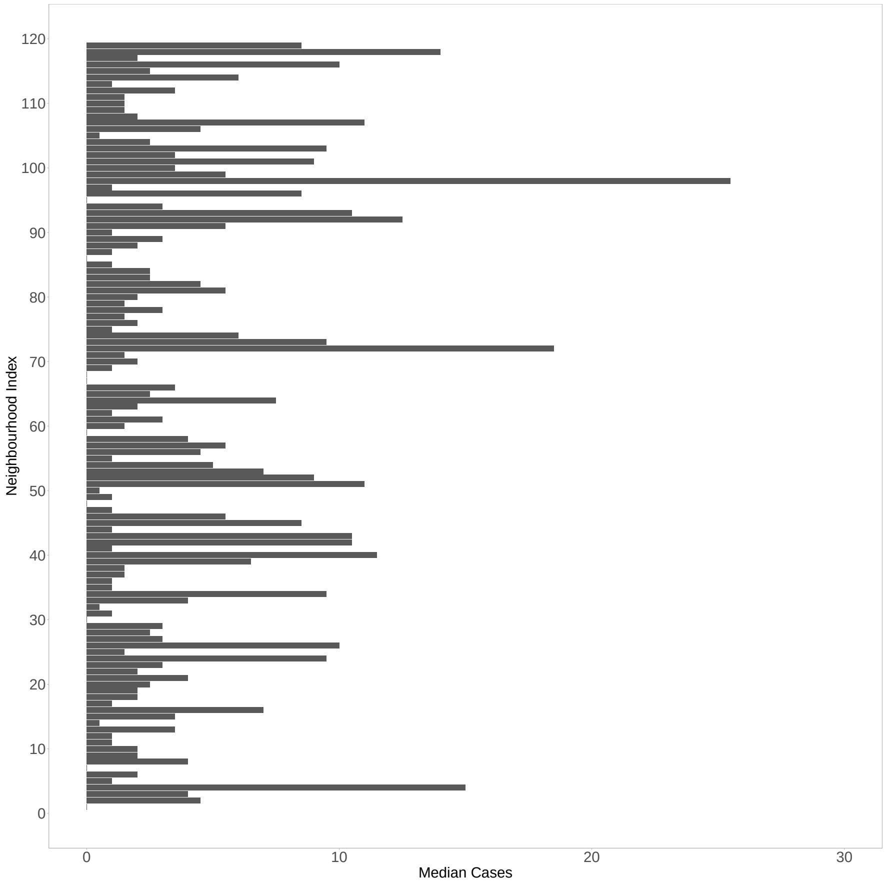


Figure SI 12: **Median weekly cases by neighbourhood during 2013 season:** For each neighbourhood, we examined the reported incident cases during a subset (July–August 2013 inclusive) of the abnormal, low-incidence season of 2013, as we had estimated lower relative contributions of high-mobility neighbourhoods during this time period. Neighbourhoods are ordered in ascending order from lowest to highest total mobility flows.
